# School start times and academic achievement - a systematic review on grades and test scores

**DOI:** 10.1101/2021.05.19.21252346

**Authors:** Anna M. Biller, Karin Meissner, Eva C. Winnebeck, Giulia Zerbini

## Abstract

School start times have been at the centre of many scientific and political debates given the accumulating evidence that bell times are generally too early, and thus lead to an epidemic of sleep restriction in the student population. Recent media attention has conveyed the message that later school starts not only improve sleep but also result in better academic achievement. Several studies have been recently published on this topic requiring a comprehensive review of the results to clarify the relationship between later school start times and academic achievement to inform the general public and policy makers.

To this end, we conducted a systematic review of the current literature on school starting times and academic achievement in middle and high school students, considering grades and standardised test scores as achievement measures. We followed the PRISMA guidelines for searching, including, and reporting relevant literature and identified 21 studies for detailed analysis. Evidence quality of included studies was assessed with a pre-defined risk of bias assessment using modified items from the GRADE scheme and ROBINS-I tool.

About half of the reviewed studies reported no (positive or negative) effect of delaying school times on grades and test scores, while the other half reported either mixed or positive results. Given the strong heterogeneity of included studies, we grouped them according to various characteristics, such as academic outcomes, dose of delay, evidence quality, or study design to identify potential hidden effects. Despite this, we could not identify any generalisable effect beyond single studies as to whether delaying school times has clear beneficial effects on academic performance.

Given that grades and scores determine future career trajectories and predict future success, the question whether school start times contribute to academic achievement is of great interest for the general public and needs to be further clarified. Mechanistically, it is very likely that improved sleep leads to or mediates improved cognitive performance and learning, but definitive conclusions on whether this also translates into better grades and scores across all students requires better evidence at this stage. Importantly, this does not preclude other positive outcomes of later start times such as improved sleep (quality), motivation or learning but draws attention on current gaps and shortcomings. To this end, we also highlight critical methodological aspects and provide suggestions to increase the evidence-level and to guide the direction of research in future studies.

## Introduction

Early school start times (SSTs) have been recognized as one of the leading causes of inadequate sleep in teenagers worldwide. They clash with the longer and later sleep needs of teenagers^e.g. 1–4^, leading to wide-spread, chronic sleep restrictions in the student population^e.g. 5–8^. Because of the accumulating evidence that sleep restriction is detrimental for psychological^9–11^ and physical health^12,13^, some schools (mainly in the US) have delayed their SSTs during the past decades.

Several studies - although mostly short-term and cross-sectional - have documented the beneficial effects of delaying SSTs on sleep duration and daytime sleepiness (as reviewed in^14–16^). More recently, other outcomes with regards to SSTs have been investigated, such as cognitive and academic performance. Since short sleep has been linked to detrimental effects on learning, memory, and cognition^17–23^, it is fair to hypothesize that delaying SSTs could result in better academic achievement (e.g. as measured in grades or scores) mediated by longer sleep duration, improved sleep quality or better circadian alignment.

However, early findings from field studies on this topic are very heterogenous, likely due to methodological differences in outcome variables and study designs^24,25^. For instance, academic achievement has been operationalised in different ways (e.g. self-reported grades, single final grades, grade point averages, standardised test scores) and with different scales. In addition, study designs vary considerably across studies, and achievement is influenced by many student- and school-level factors^e.g.26–30^.

Previous reviews have mostly summarized the effects of delaying SSTs on several different variables (e.g., sleep, tardiness rates, absences, motor vehicle accidents and health^14,16,25^). We identified a total of 12 peer-reviewed reviews^15,16,37,38,24,25,31–36^ – only 3 of them systematic reviews^15,16,34^ – that discuss SSTs in relation to academic achievement. However, all of them cover the topic only broadly or on the side, so that no unifying conclusion can be drawn from the existing reviews to date. Despite this lack in systematic reviews and meta analyses, newspaper articles often purport it as established scientific fact that later SSTs improve academic achievement^39–41^, while some public outreach programs also convey this message^42^, mostly referring to single studies that found positive associations.

Since academic achievement shapes future career trajectories^43–45^, answering the question whether delaying SSTs improves achievement goes beyond simple and genuine scientific curiosity - a rigorous and up-to-date analysis of the accumulating evidence is warranted. Following the PRISMA guidelines for systematic reviews and including a detailed risk-of-bias assessment based on items from the GRADE scheme^46^ and the ROBINS-I tool^47^, we addressed the specific gaps in the review literature to date, such as a particular need for discussion of the quality of evidence, a detailed description of the outcome variables and statistical analyses, and a distinction between middle/high school and college students, who differ considerably in their sleep characteristics and class schedules. In our review, we thus systematically assessed the existing evidence on SST effects on academic achievement via studies on course grades or standardised test scores in middle and high school students. We provide both a summary as well as detailed descriptions of each included study, assess the overall and individual evidence level and highlight critical points for future research.

## Methods and Materials

### Literature search

Our focused question was whether changes in school start times in middle or high schools (or international equivalents) have any effect on academic achievement as measured in (standardised) test scores or course grades (both subjectively and objectively reported). Therefore, we conducted a systematic electronic literature search in Web of Science and PubMed via Endnote (version 9.3.1), and an online search on SCOPUS in August 2020, which was updated in November 2020. All languages, article types or year of publications were allowed. The following search terms were used (in title, abstract or keywords):

*school start times* OR *school start time* OR *school starting times* OR *school start delay* OR *start late* OR *start early*

AND

*grades* OR *school performance* OR *academic performance* OR *test scores* OR *standardised scores* OR *achievement*

Additionally, reference lists of previous reviews and articles were scanned to ensure complete retrieval. We included two unpublished articles that are currently under review in peer-reviewed journals^48,49^. The PRISMA flowchart (Fig. 1) was followed to adhere to preferred reporting guidelines for systematic reviews^50^.

**Fig. 1.**
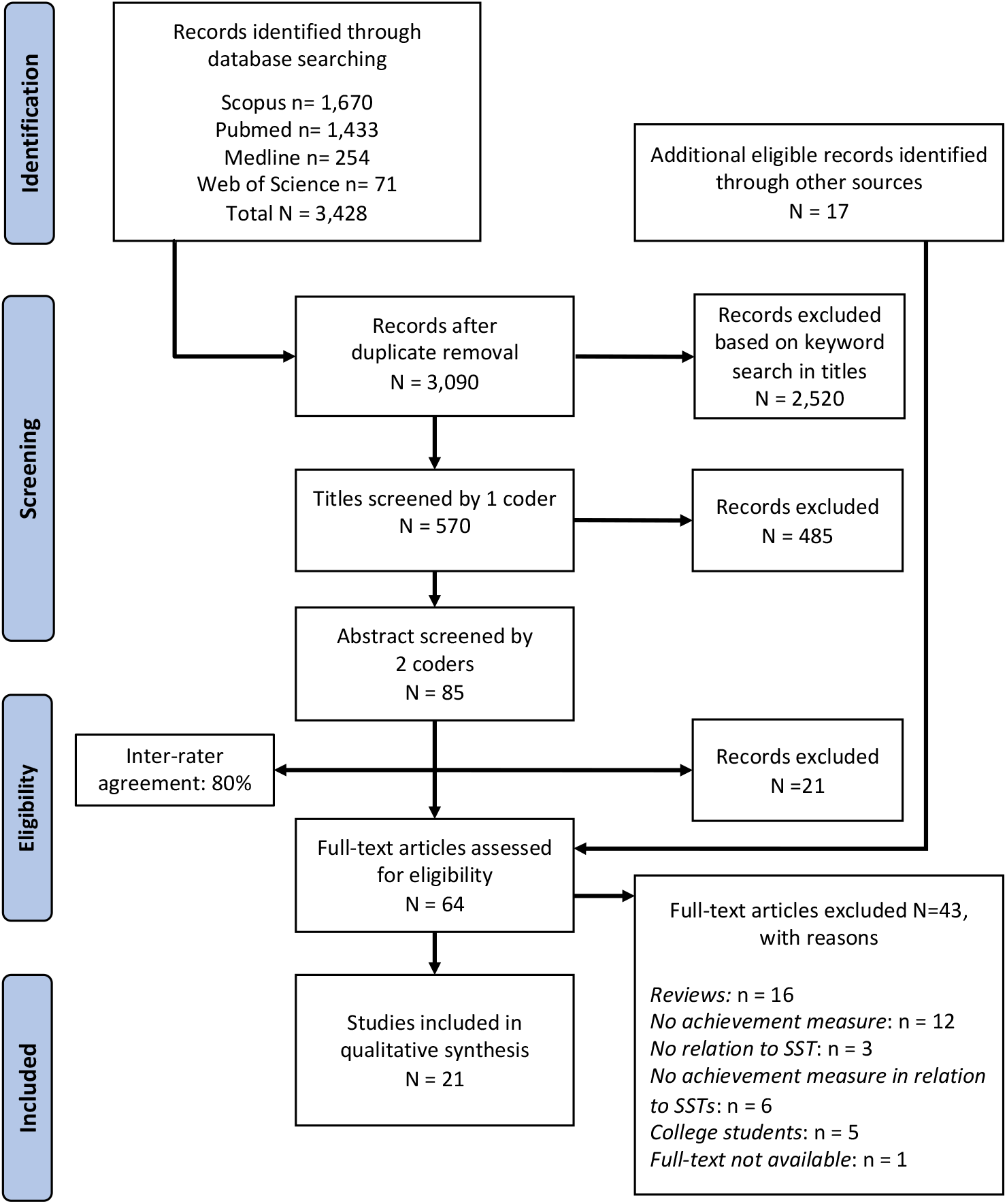
PRISMA flowchart. The PRISMA flow diagram for our systematic review process detailing the database searches, the number of identified records, titles and abstracts screened, the final studies included in qualitative synthesis and reasons for exclusion of studies.

### Study selection criteria

All duplicate retrievals were removed via the Endnote duplicate function followed by a manual search and deletion round. All remaining titles and abstracts were subsequently screened on relevance with regards to the focused question. Full articles were then searched if the following study selection criteria were fulfilled: academic achievement was assessed as grades or (standardised) test scores; participants were middle school or high school students; articles included both a change/variation in SSTs and a measure of academic achievement (course grades or standardised test scores).

### Data abstraction and analysis

The recommended PRISMA guidelines for data synthesis and systematic reviews were followed^50^. AMB and GZ independently and systematically extracted pre-defined study characteristics as per Figure 1. The 21 studies included in the final qualitative synthesis were grouped by their study design. Please note that we grouped according to the design underlying the grade or test score outcomes, which can differ from the design for other outcomes investigated in the respective study such as sleep duration. Identified designs were: longitudinal designs with control group, longitudinal designs without control group, and cross-sectional designs. Note that a longitudinal design means that the *same students* were followed over several time points (within-subject comparisons) whereas a cross-sectional design compares *different students* either at one time point or between time points (between-subject comparisons). It was noticed that several cross-sectional studies described their design as longitudinal because they followed the same *schools or districts* over several time points (which might or might not include the same students); we considered these studies as *repeated cross-sectional studies* based on their study design and statistical analysis. Authors were contacted when information was missing, not clearly defined or further analyses were available upon request. If authors responded, information was updated accordingly. If authors did not answer or failed to provide necessary information in the original article, this was marked as “not available” (“NA”) in Tab. S1 and, and flagged orange or red (depending on the severity) in the reporting bias category of the risk of bias assessment (Tab. 1).

**Tab. 1.**
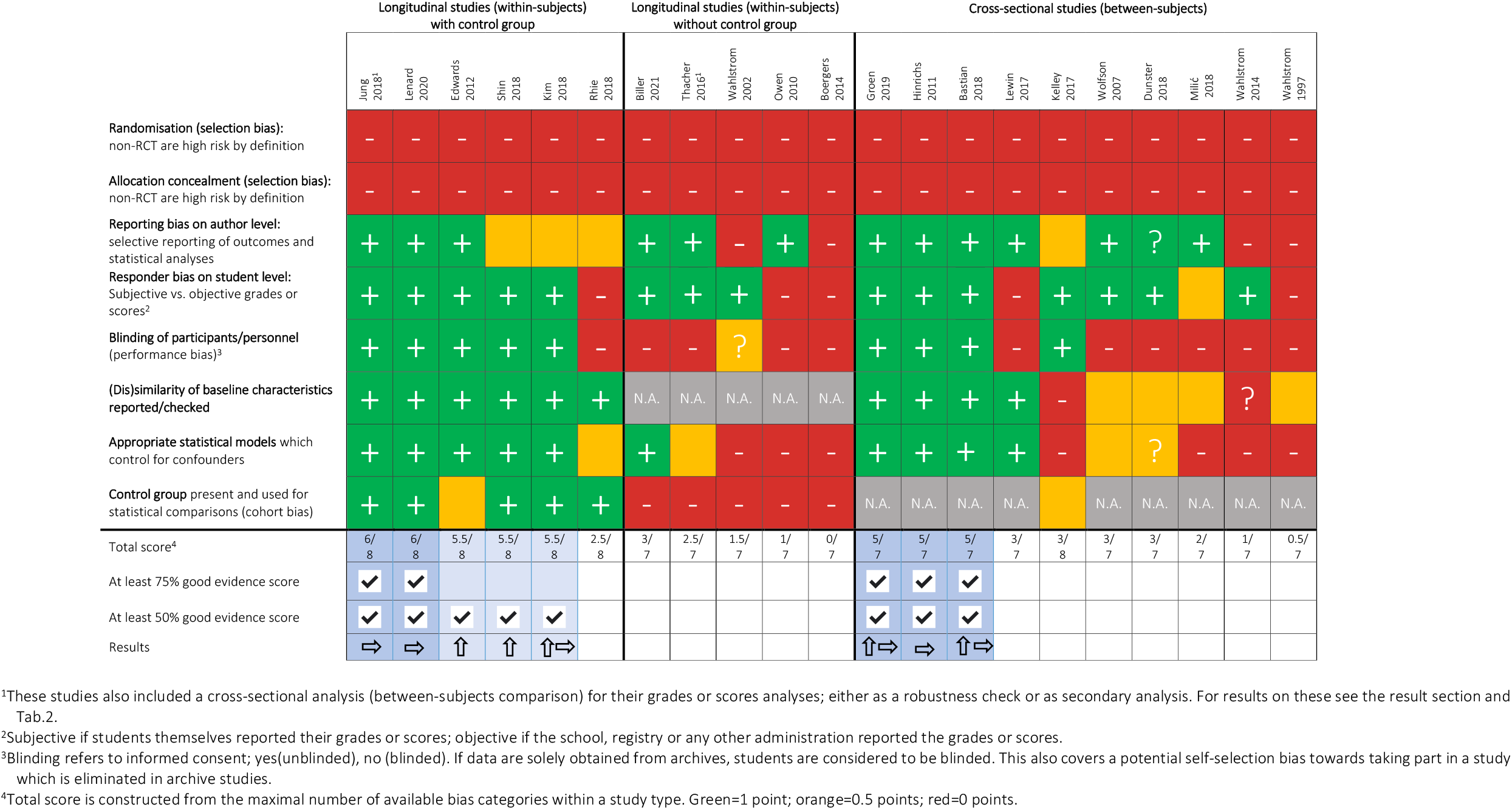
Risk of bias assessment. Included studies are ordered by their study design (used for grade or score analyses) and assessed in different bias categories. Cell colour shows the risk status for the respective bias category (red=high risk; orange=intermediate; green=low risk). Question marks indicate ambiguous information (more details given in Tab. S1). For the final study result based on the obtained evidence score, an upward arrow indicates a positive finding for later school start times on academic achievement, a right arrow indicates mixed findings. Longitudinal studies assess the same individual over time, while cross-sectional studies follow different students. NA, not applicable.

### Risk of bias assessment

A pre-defined risk of bias assessment was conducted independently by AMB and GZ (Tab 1). Given that there were no randomised controlled trials (RCTs) in the final sample and the large methodological differences between studies, bias assessment guidelines were adapted as there are no standard guidelines for non-RTCs. To this end, items from the GRADE scheme^46^ and ROBINS-I tool^47^ used for non-RTCs were included and modified. Each study was evaluated on the following bias categories and flagged green (low risk), orange (intermediate risk) or red (high risk):

#### Selection bias (randomisation)

Participants were not randomly assigned to the control group or the treatment group. Non-RTC are high risk by definition.

#### Allocation concealment

Researchers did not know the sequence or method of randomisation and hence could not predict the next allocation. Non-RTC are high risk by definition.

#### Reporting bias on author level

Authors did not report or only partially reported all outcome variables, sources of outcomes, statistical analyses or general information necessary to judge the study. When a publication stated that information was available upon request, the authors were contacted.

#### Responder bias on student level

Students could be biased when self-reporting, which is not the case for objectively reported grades or scores provided by official sources (e.g. the registry or state level administrations).

#### Performance bias (blinding of participants/personnel)

Participants who knew that they took part in a study are prone to behavioural changes (Hawthorne effect). If informed consent was obtained, students were considered unblinded, else they were blinded. This also covers a potential self-selection bias towards taking part in a study.

#### (Dis)similarity of baseline characteristics

Authors checked and reported the (dis)similarity of baseline characteristics between cross-sectional groups or between control and treatment groups.

#### Appropriate statistical models

Statistical analyses accounted for confounders and were appropriate for the given study design.

#### Cohort bias (control group present)

Longitudinal changes might be due to cohort characteristics and not due to an intervention when no control group was present. Only applies to longitudinal studies.

Tab. S1 lists the decision criteria underlying the risk of bias assessment. In cases where the assessment differed between AMB and GZ, mutual agreement was sought after discussion of critical points. In case no agreement could be reached, two independent scorers (ECW and KM) evaluated the respective studies and a consensus was found across all scorers. From the assessment, a total quality-of-evidence score was calculated as follows: scores for each bias category were added up (green contributed 1 point, orange 0.5 points and red 0 points) and then divided by the maximal possible score (8 for the longitudinal studies with control group, 7 for the longitudinal studies without control group, and 7 points for cross-sectional studies). The quality-of-evidence score was the proportion (%) of the maximum score (e.g. 6 out of max 8 points = 75%). The different bias categories were not weighted. We defined scores <25% as low, ≥25% and <75% as moderate and ≥75% as good.

## Results

### Literature search

A total of 3,428 articles were identified based on the automated search in title, abstract and keywords, of which 3,090 remained after duplicate removal (Fig. 1). Due to this large number, a second automated search was carried out on titles only, resulting in 570 articles. One coder (AMB) then screened titles excluding 485 manually due to irrelevant titles. The abstracts of the remaining 85 studies were screened by both coders (AMB and GZ), who agreed on 47 studies (80% inter-rater agreement) and additionally identified 17 studies through reference lists of included studies. The identified 64 articles were subsequently screened in full by both coders, 43 excluded based on the pre-defined exclusion criteria and 21 ultimately included in the qualitative synthesis (Fig. 1).

### Study characteristics and quality

In the following paragraphs, summary information concerning all included studies are reported (see also Fig. 2 and Tab. 2).

**Fig. 2.**
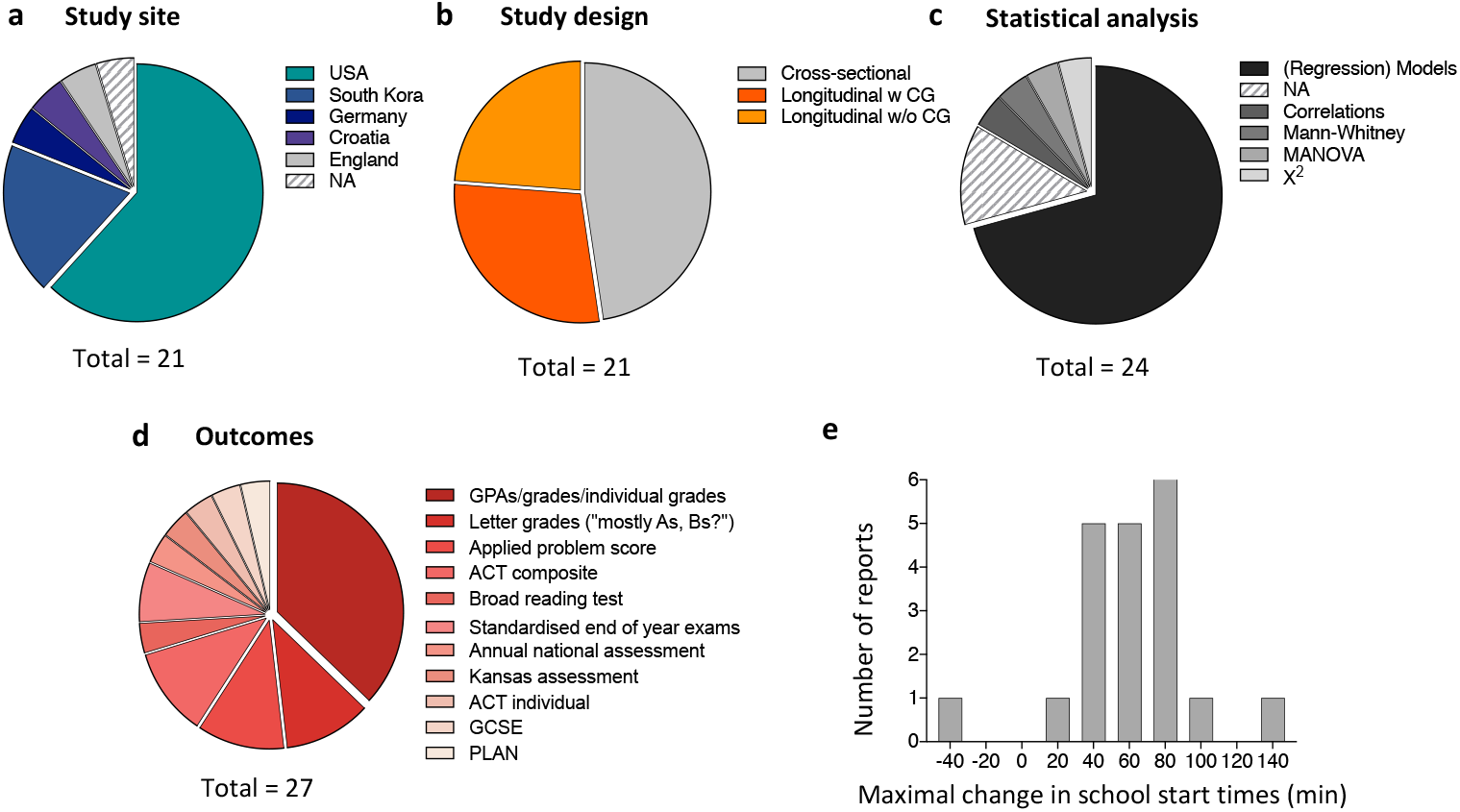
Characteristics of included studies. **a-d**, Pie charts depicting key characteristics of the 21 studies included in the final review. Since several studies used multiple types of analysis or assessed multiple outcomes, the total number in c,d is >21. **e**, Histogram displaying the magnitude of the school start changes reported in the 21 studies. When a study reported ranges, the maximum of the range was taken. Please note that these numbers therefore just provide a rough overview and are not precise. Abbreviations: NA, not available; w, with; w/o, without; CG, control group; GPAs, grade point average; ACT, American College Test; GCSE, General Certificate of Secondary Education; PLAN, a preliminary ACT test discontinued in 2014.

**Tab. 2.**
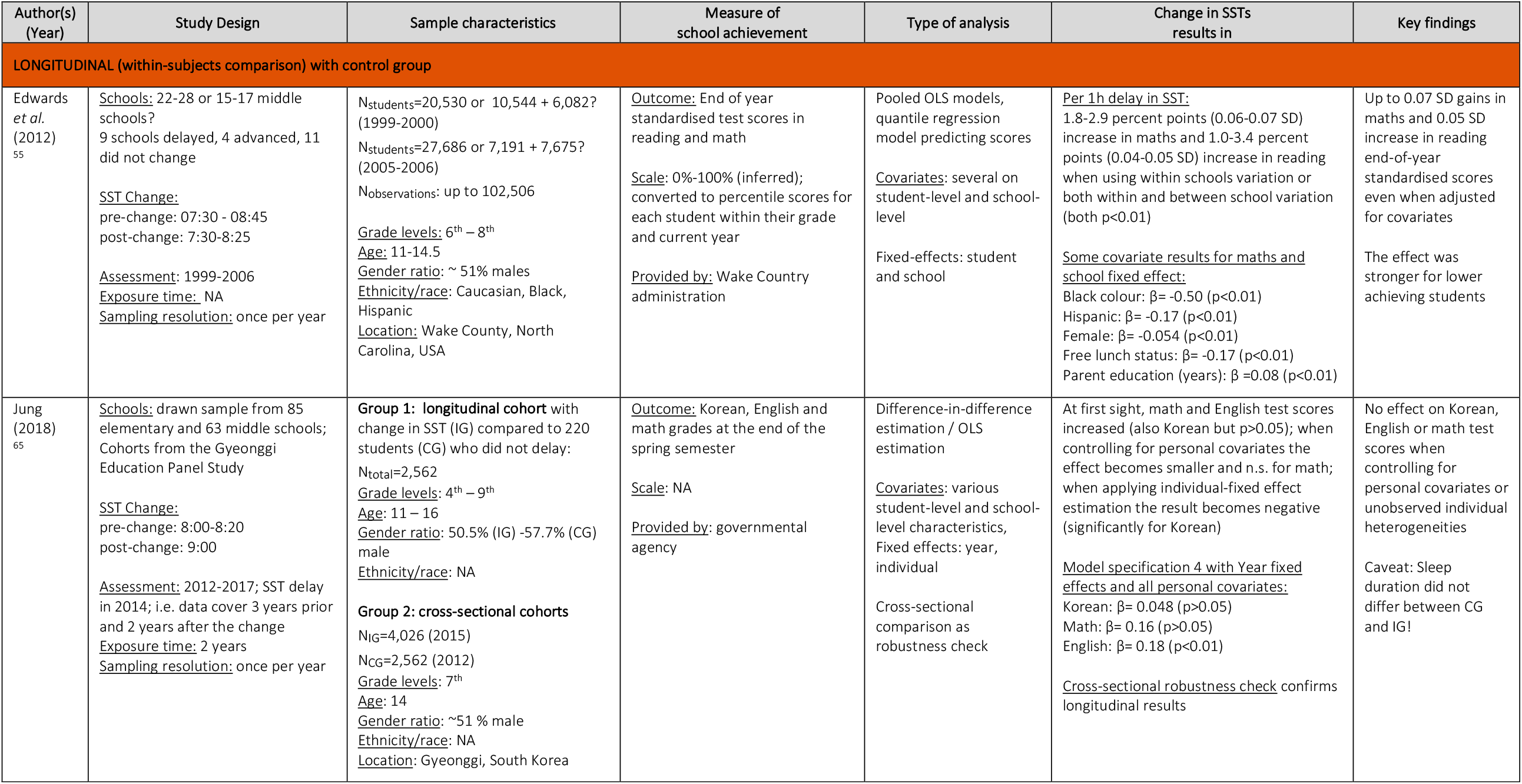
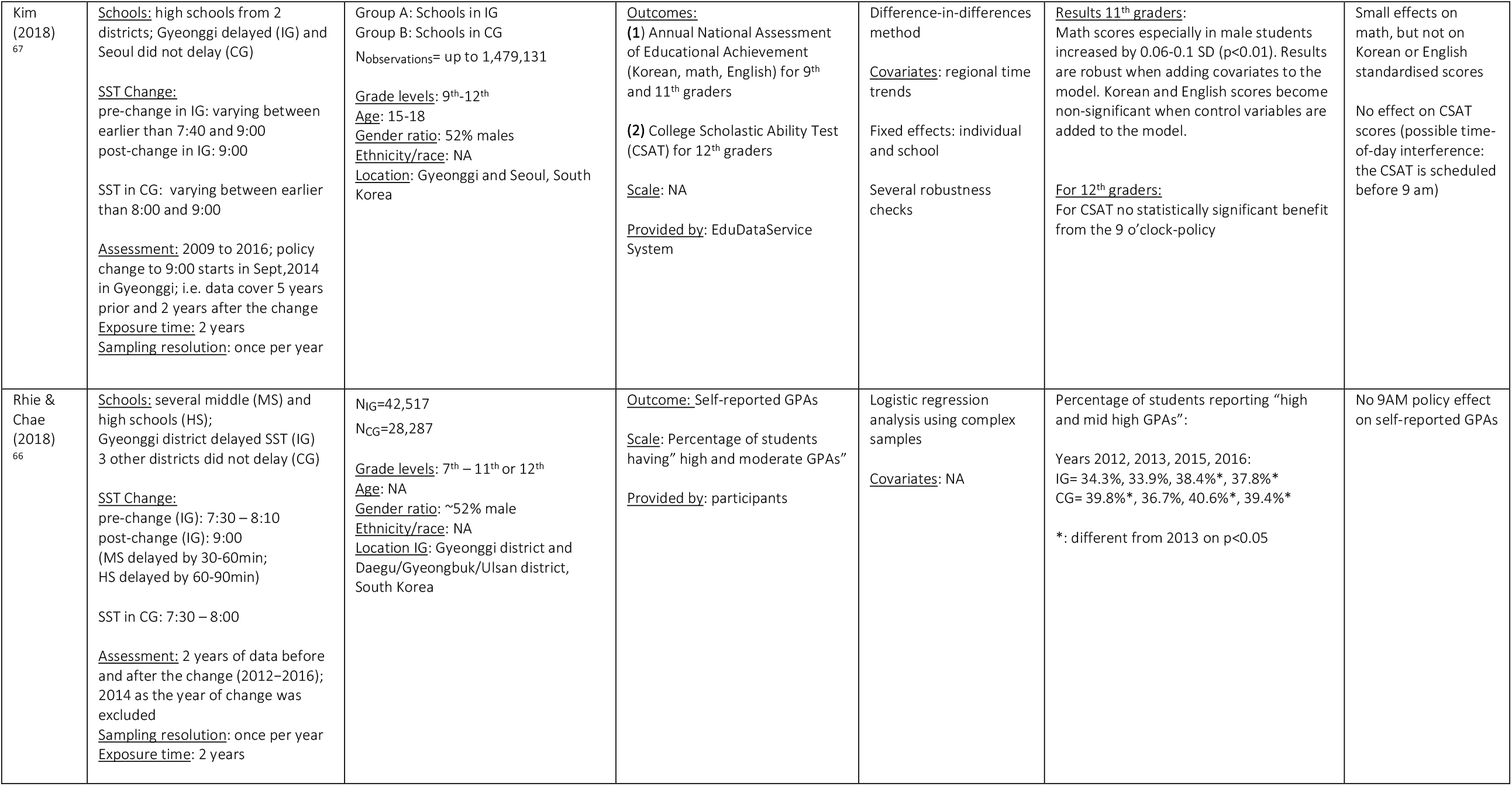
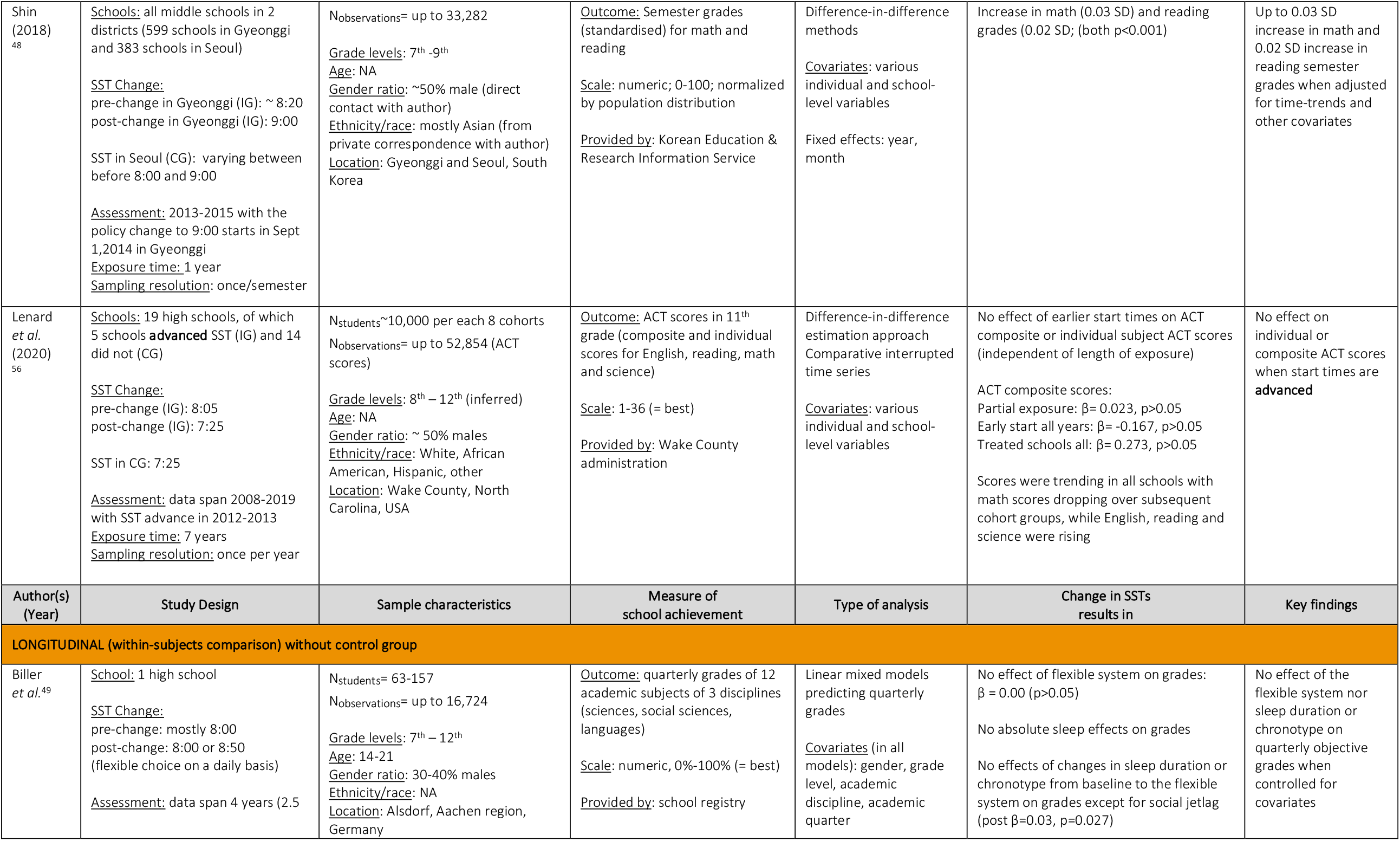
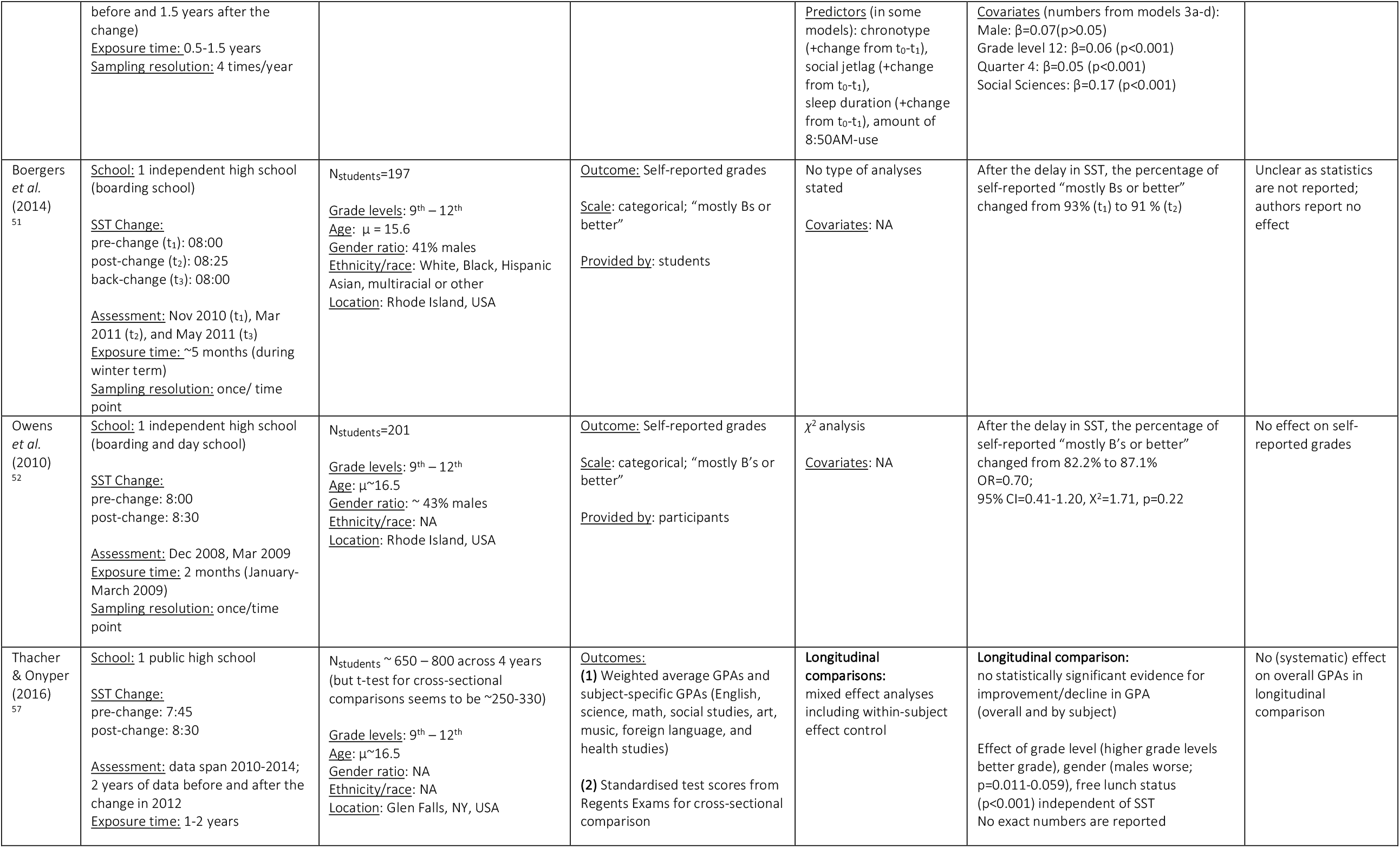
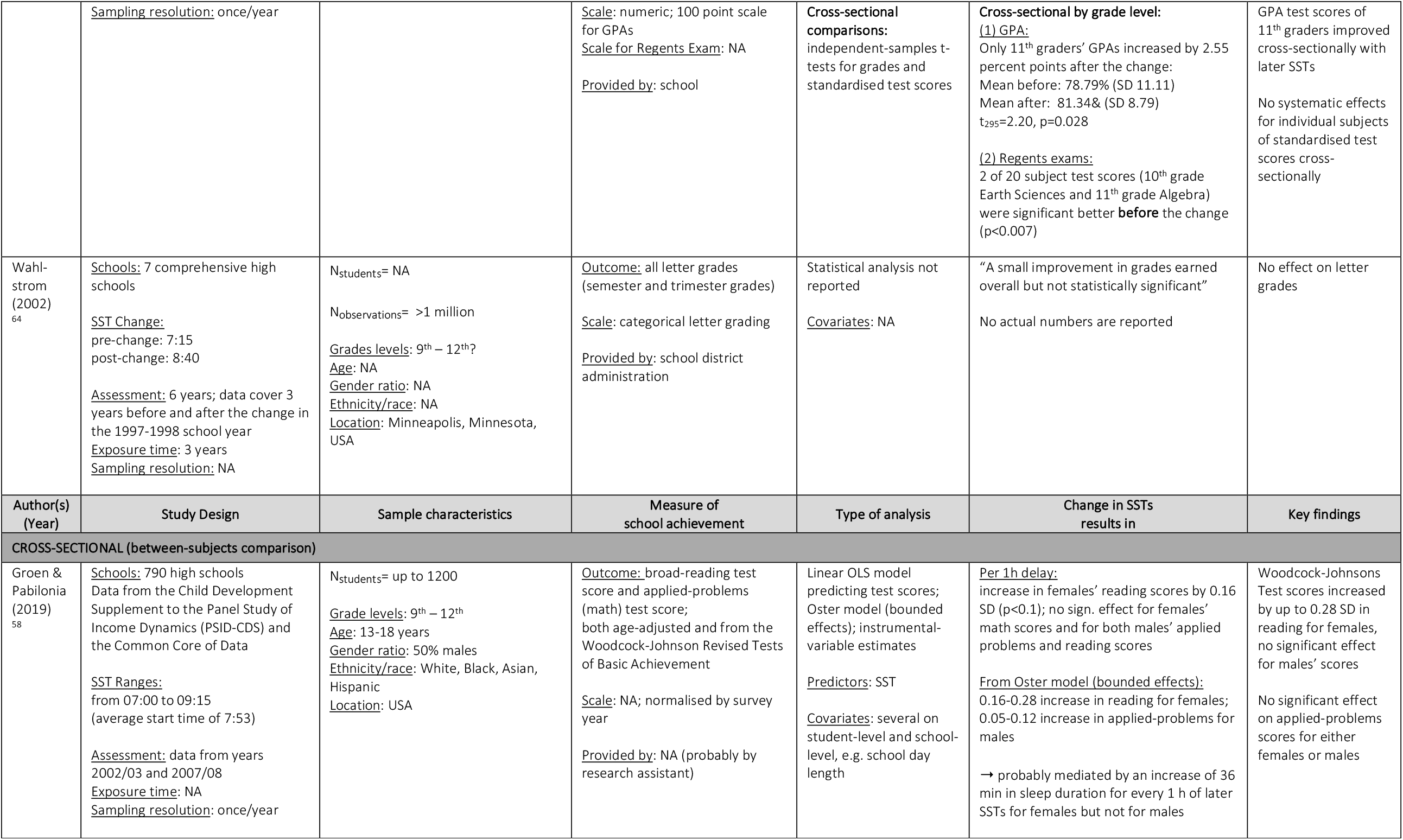
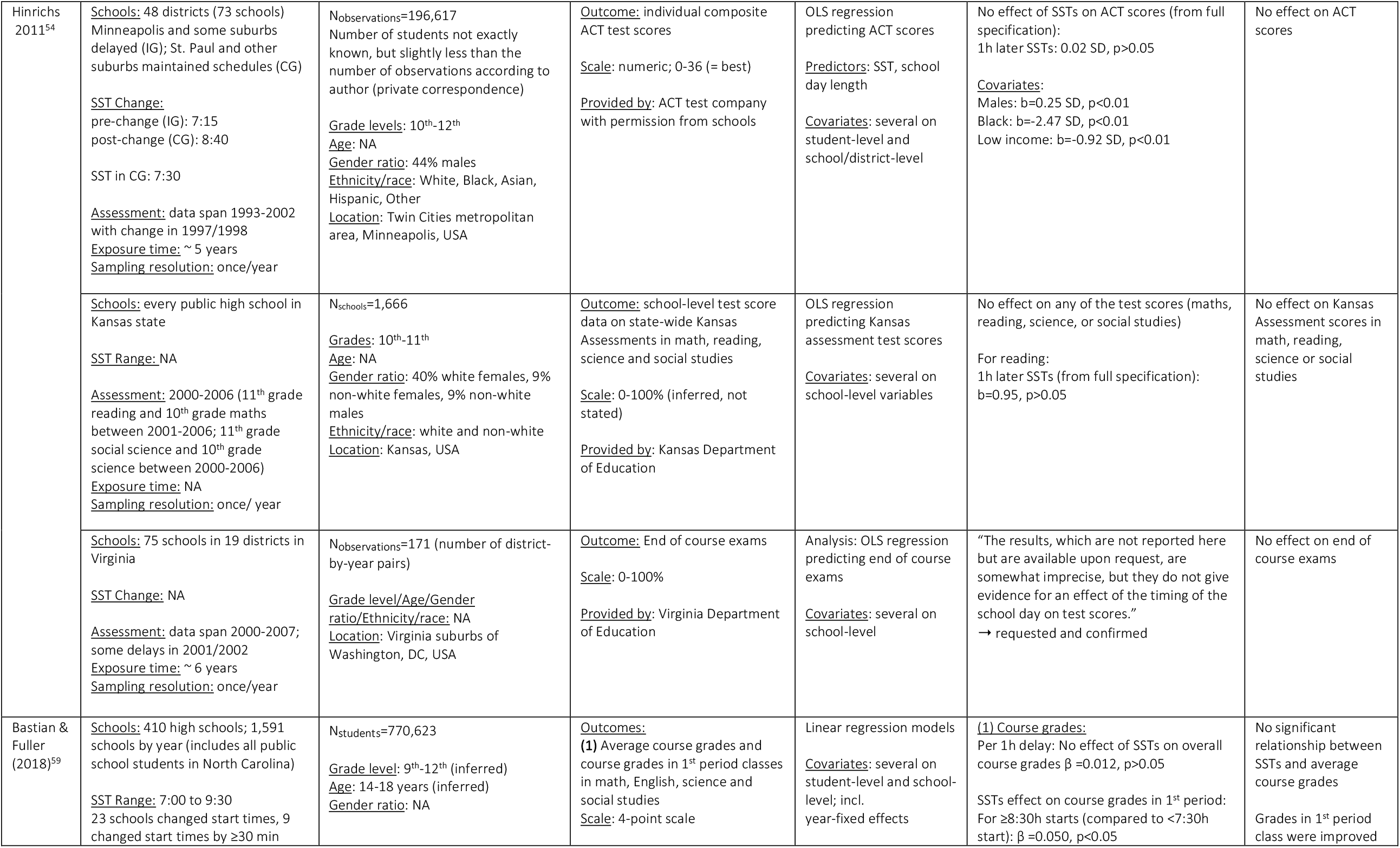
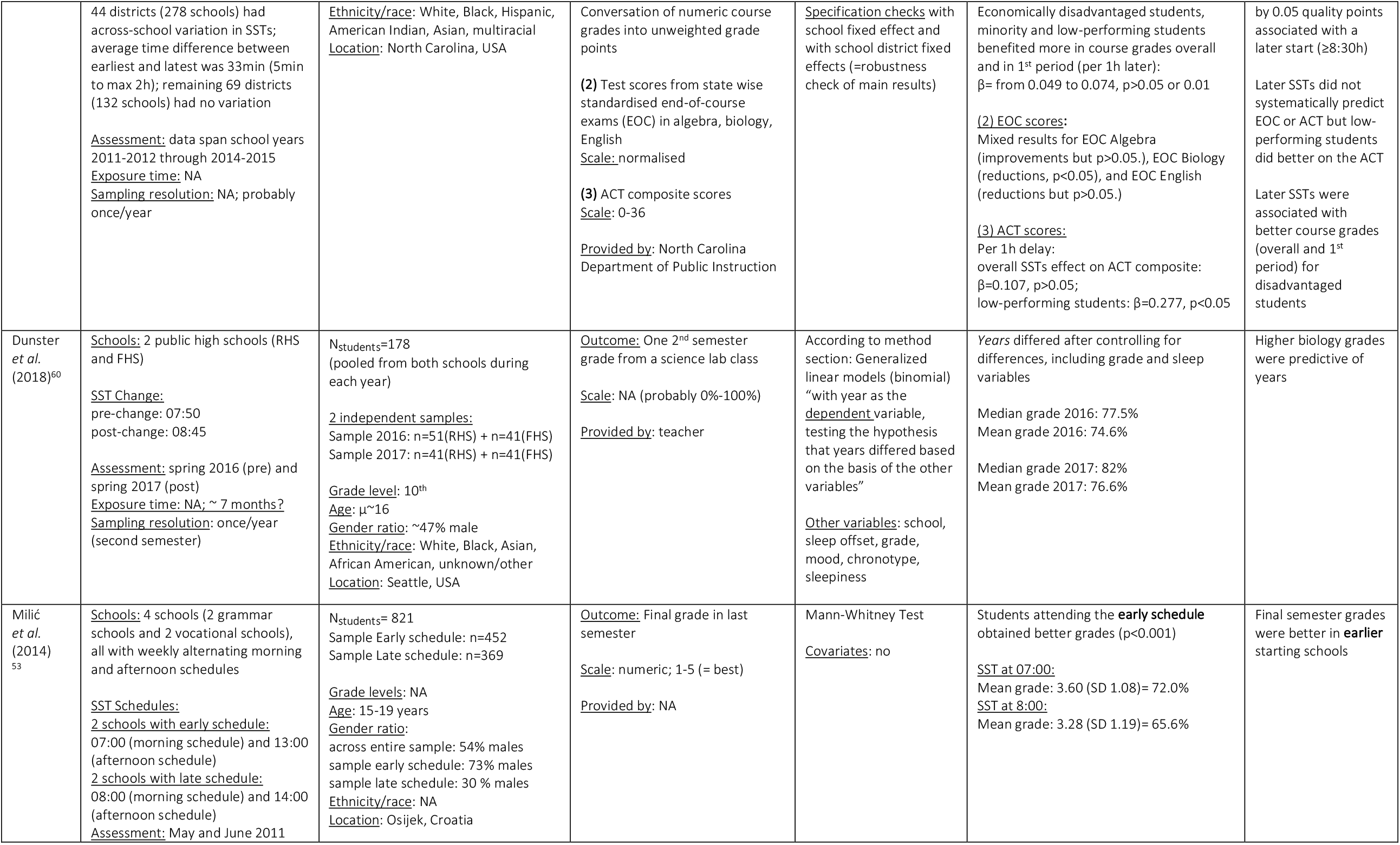
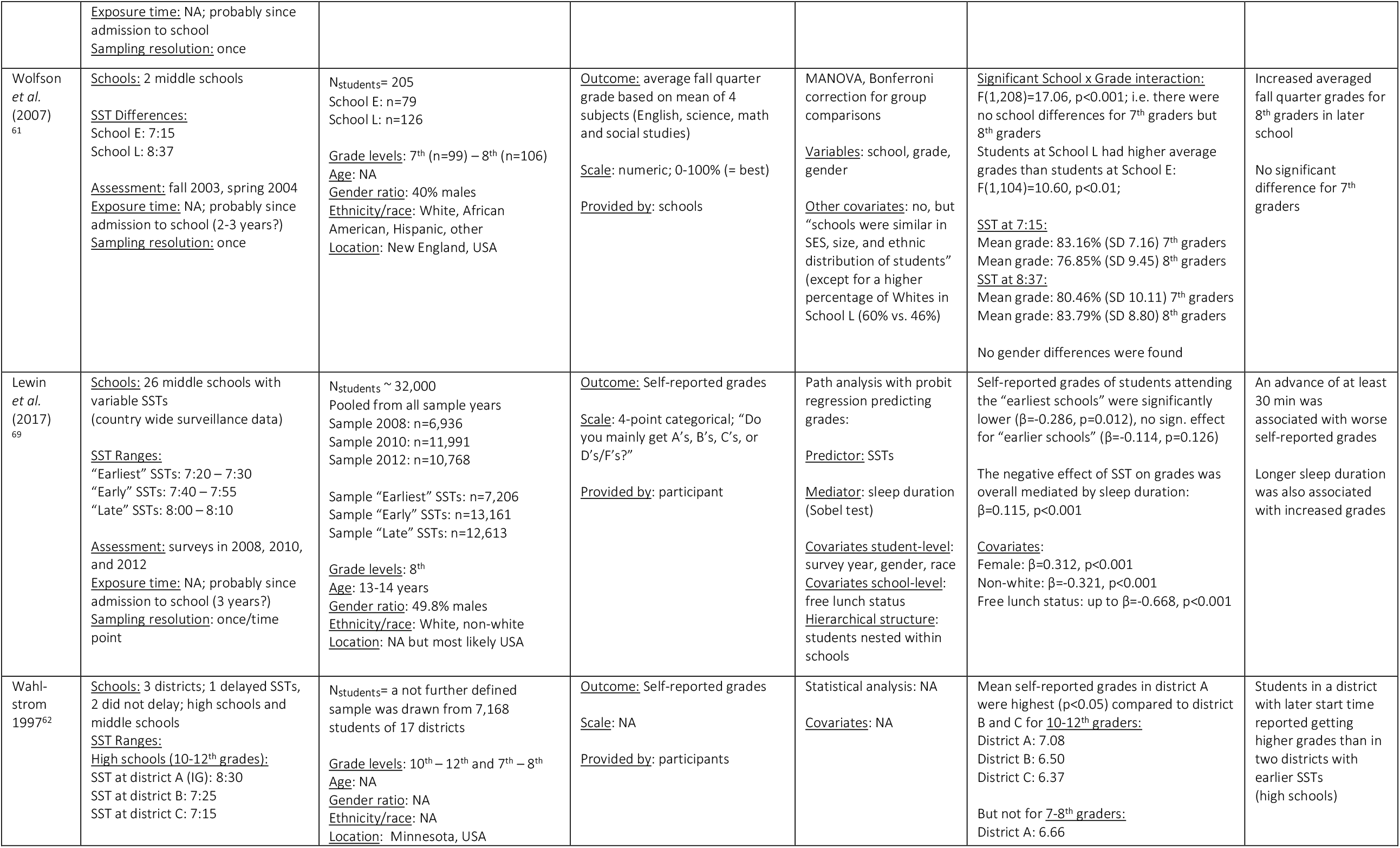
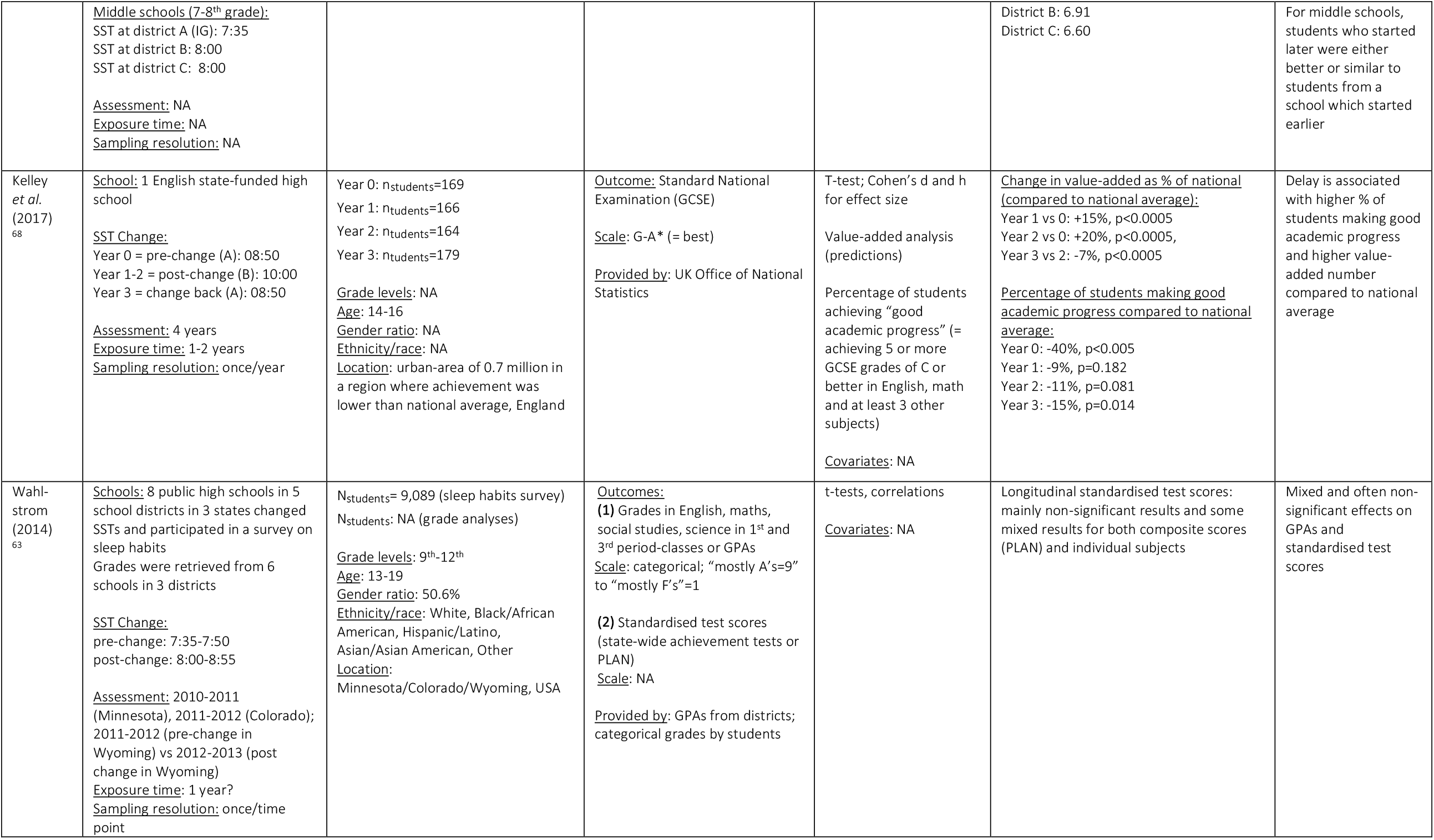
Detailed descriptions of included studies. Studies are ordered by their type of study design (longitudinal with control group, without control group and cross-sectional). Study designs of two studies could not clearly be clearly defined. Abbreviations: SST, school start time; OLS, ordinary least square; SD, standard deviation; b, unstandardised beta coefficient; β, standardised beta coefficient; µ, average; CG, control group; IG, intervention group; CSAT, College Scholastic Ability Test; GPA, grade point average; ACT, American College Test; OR, odds ratio; NA, not available.

#### School type and cohort characteristics

The majority of studies collected data in high schools (>900 schools), of which 2 were also boarding-schools^51,52^, 2 grammar schools and 2 vocational schools^53^. Other school types were middle schools (>140) and elementary schools (85, not considered here). In one study, school type was not specified^54^. The sample sizes varied drastically between 157 to >770,000 individual students and up to >1 Mio number of observations (e.g. individual grades). However, some authors did not distinguish between number of individuals, number of schools and number of observations. In 13 studies, age of participants was reported and ranged approximately between 11-19. Most studies were conducted in the US (13)^51,52,62–64,54–61^, followed by South Korea (4)^48,65–67^, Germany^49^, Croatia^53^, England^68^, and one unknown location^69^ (Fig. 2a and Tab. 2). Gender ratios, ethnicity/race and a proxy for socioeconomic status (SES; free or reduced lunch eligibility) were not consistently reported.

#### Study designs

We identified longitudinal (within-subject) and cross-sectional (between-subject) studies. The 11 longitudinal studies all included a change in SSTs and hence had an intervention group^48,49,67,51,52,55–57,64–66^. However, only 6 studies had an additional control group with no change^48,56,65–67^ or advance of SSTs^55^ (Fig. 2b). Of the cross-sectional studies, 4 studies compared schools in various districts without an intervention but based on their different school start times^53,58,61,69^. The rest included a change in SST, providing mostly repeated cross-sectional comparisons of schools or districts over roughly one^60^ or several years^54,59^, or at one time point after the change^62,63^. One cross-sectional study also had an A-B-A design, in which the school start delay during phase B was abolished to return to baseline start time(A) after 2 years^68^.

#### Statistical analyses

A vast range of different statistical analyses was reported (Tab. S1 and Fig. 2c). Notably, regressions were the dominant analysis method, ranging from general OLS regressions^54,55,58,59,65^, quantile regression^55^, difference-in-difference methods^48,56,65,67^ and binomial regression^60,66^ to linear mixed models^49,57^ and path analysis with probit regression^69^. One study reported Oster models with bounded effects and instrumental estimates^58^. Another study used MANOVA^61^, while several simpler analysis methods not controlling for covariates were also used. These were t-tests^57,63,68^, X^2^-tests^52^, Mann-Whitney Test^53^ and correlations^63^. Notably, several studies did not report statistical analyses^51,62,64^.

#### Study outcome measures

About half of the studies provided grades as outcome measures, while the other half provided (standardised) test scores. However, since some studies did not provide explanations whether scores originated from standardised tests, a clear distinction between course grades and test scores was not always possible. Clearly defined scores were ACT scores (American College Test)^54,56,59^, national achievement scores or PLAN scores^63^, standardised test scores from Regents Exams^57^, standardised end-of-course exams^59^, annual national assessment of achievement in South Korea^67^, GCSE in the UK (General Certificate of Secondary Education)^68^, and Woodcock-Johnson Revised Test of Basic Achievement scores^58^, all of which were objectively reported (except for Groen *et al*., for which the source was unclear^58^) (Fig. 2d). The remaining studies analysed other types of objective scores or grades ^48,49,55,59,61,64,65^, subjective grades^51,52,60,62,63,66,69^, and in one study the outcome was unclear^53^. Sampling resolution was mostly once per year, the highest reported resolution was once per academic quarter^49^.

#### Amount of school start time change and duration of exposure to the new start time

The SST delay reported was on average 64min (median=60, SD=26) with a range of 25-135 min (Fig. 2e). This average is based on the maximal delay reported by each study (several reported multiple amounts) and thus an approximation. Since some studies only provided SST ranges or a minimal start delay, the numbers are not precise. In 2 studies, SSTs were actually advanced by 40 min and 25-45 min respectively^55,56^. One study changed to a flexible SSTs in which students could choose daily whether to attend school at 8:00h or 8:50h^49^. Exposure duration to the (new) start time ranged from as little as 3 months to 7 years (Tab. 2). However, several studies did not clearly state the timeframe (so we inferred where possible), or did not test a change but a difference in start times across schools.

### Summary of individual study results

In the following, we shortly report findings of individual studies, grouped by study design. In summary, 5 studies found clear positive effects of delayed school starts on academic achievement^48,55,62,68,69^, 5 reported mixed effects^58,59,61,63,67^, 9 did not detect significant effects^49,51,52,54,56,57,64–66^, 1 reported negative effects^53^, and one study’s finding was unclear^60^ (Fig. 3b). Regarding SST *advances*, 1 study reported negative effects^69^, while another did not^56^.

**Fig. 3.**
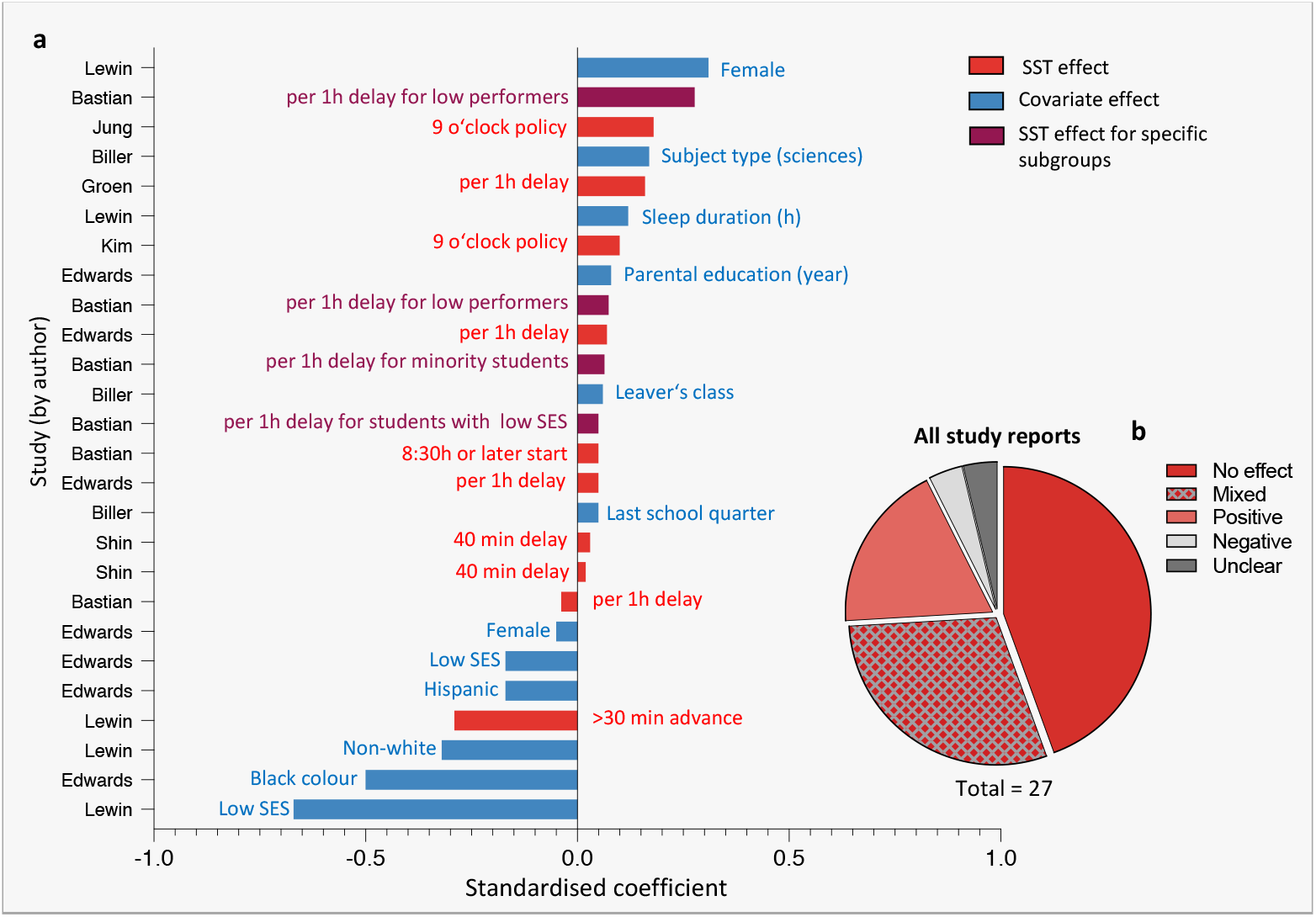
Overall study results and effects sizes. **a**, Standardised beta coefficients ordered by magnitude and study author from the subset of studies that reported standardised coefficients and statistically significant effects (n=8 of 21). Only these statistically significant effects are depicted, non-significant ones were left out. Standardised coefficients are in units of standard deviation of the outcome variable. Quarter refers to the academic quarter of a school year in Germany. Low socioeconomic status was measured as free lunch status. For exact study references see Tab. 1. **b**, Summary of simplified findings from all included studies (N=21). The total is >21 since several studies reported multiple outcomes. Abbreviations: SST, school start time; SES, socio-economic status.

Notably, of the 21 studies, 4 studies investigated the same 9 o’clock policy in South Korea^48,65–67^. Although they considered partly different outcomes and schools (middle vs high schools), the Korean studies likely analysed data from overlapping students, hence this cannot be regarded as entirely independent evidence. The same may apply to 2 studies by Wahlstrom *et al*. conducted in the same district: the report in 2002^64^ might be a longitudinal follow-up of the report from 1997^62^, but we were unable to confirm this.

#### Longitudinal studies with control group

**Edwards (2012)^55^** followed several middle schools in Wake County, North Carolina (USA), over 8 years (up to N_observations_>102,000) during which 9 schools delayed, 4 advanced and 11 did not change their SSTs. The authors analysed objective standardised end-of-year test scores in reading and math via regression models with pooled OLS models and accounted for various covariates both on the student and school level. They found that a 1h later school start corresponded to a 1.8-2.9 percentile increase in math (0.06-0.07 SD) and 1.0-3.6 increase in reading (0.04-0.05 SD) when adjusted for covariates (both significant), and that the effect was stronger for lower achieving students.

**Jung (2018)^65^** followed 85 elementary and 63 middle schools (N_students_>4,000) in South Korea 3 years prior to and 2 years after a delay from 8:00h-8:20h to 9:00h. Participants were recruited as part of the Gyeonggi Education Panel Study and their objective Korean, English and math course grades were reported. The author found no effect of delaying SSTs on grades in the longitudinal within-subject comparison with the control group (difference-in-difference estimation/OLS estimation). Cross-sectional analyses as robustness check confirmed the longitudinal results. Similar to Kim ^67^ and Biller *et al*.^49^, the author also found that when not controlling for covariates, test scores increased, while the effect became statistically non-significant when covariates were added.

**Kim (2018)^67^** also compared high schools from two districts in South Korea (N_students_>2,000), of which Gyeonggi adopted a 9 o’clock start time policy. Pre-change SSTs in this district ranged from 7:40h-9:00h and were delayed to 9:00h post-change, while Seoul did not change (control group). The author used the difference-in-difference method and mixed within-between regression models to estimate the influence of the 9 o’clock policy on the objective Annual National Assessment of Educational Achievement for 9^th^ and 11^th^ graders, and the College Scholastic Ability Test (CSAT) for 12^th^ graders (data cover 5 years pre and 2 years after the change). Only male 11^th^ graders showed a significant increase of 0.06-0.08 SD for math, even after adjusting for confounders. CSAT scores did not increase significantly with the 9 o’clock policy.

Similarly, **Rhie and Chae (2018)^66^** studied middle and high schools in 4 South Korean districts, of which Gyeonggi delayed SSTs (baseline from a range of 7:30h-8:10h) to 9:00h, while Daegu, Gyeongbuk and Ulsan did not change (SSTs ranged from 7:30h to 8:00h; control group). Based on logistic regression analysis in their large sample (N_students_>42,000), they found that self-reported GPAs increased year by year in both the intervention and the control group (data cover 2 years pre and after the change; adjustments for covariates not reported).

**Shin (2018)^48^** is the fourth study that investigated the South Korean 9 o’clock policy effects in Gyeonggi (change in SST from around 8:20h to 9:00h), compared to Seoul (control group), but the author used objective semester grades as outcome and focused on middle schools (N_observations_>33,000). The data span 2 years and was analysed using the difference-in-differenced method, which accounted for various individual and school-level variables. Shin reported a 0.03 SD increase in math and 0.02 SD increase in reading grades when adjusted for time trending (both significant).

**Lenard *et al*. (2020)^56^** looked at 19 high schools in Wake Country, North Carolina, USA, of which 5 had advanced their SSTs from 8:05h to 7:25h, while the control group (14 high schools) kept their start at 7:25h. They found no significant change in objective standardised American College Test (ACT) scores, neither in their longitudinal nor their cross-sectional comparison of about N_students_~10,000 in 8 cohorts. The authors used a difference-in-difference approach and comparative interrupted series controlling for various individual and school-level variables. Their data spanned 4 years prior and 7 years after the change.

#### Longitudinal studies without control group

**Biller *et al*. (2021)^49^** investigated in a German secondary school the effects of a unique SST change, the introduction of *flexible* SSTs, on objective, quarterly grades for up to 2.5 years prior to and 1.5 years after the change. In the flexible system, students chose daily whether to attend school at 8:00h or 8:50h after starting predominantly at 8:00h in the conventional system before. Longitudinal linear mixed model analyses of 16,724 grades in 12 academic subjects (N_students_=157) indicated that the flexible system did not affect grades when accounting for several student and school-level factors.

**Boergers *et al*. (2014)^51^** studied an independent high school (boarding school) in Rhode Island, USA, that delayed its start time from 8:00h to 8:25h (N_students_=197). The percentage of students who reported to obtain “mostly Bs or better” changed from 93% to 91% after 2 months, however statistical analyses were not reported.

**Owens *et al*. (2010)52** used the same outcome variable as Boergers *et al*.^51^ in their study of N_students_=201 from an independent high school (boarding and day school) in Rhode Island, USA, over 6 months (3 time points of assessment). They found that a school start delay from 8:00h to 8:30h was associated with a non-significant increase of students reporting to obtain “mostly Bs or better” (82% pre vs 87.1% post, using a *χ*^2^ test). Adjustment for covariates was not reported.

**Thacher & Onyper (2016)^57^** studied N_students_~800 from one public high school in Glen Falls, NY, USA, which delayed their SSTs from 7:45h to 8:30h. They used mixed effect analyses to analyse longitudinal effects (2 years before and 2 after the change), adjusting for multiple covariates and including moderator effects. This analysis indicated no systematic effect on subjectively reported GPAs (0-100%) nor subject-specific GPAs or standardised test scores (Regents exam). They did find positive effects for 11^th^ graders’ overall GPAs, however, only when they ran cross-sectional comparisons (increase from 78.79% to 81.34%). In contrast, no systematic effects on individual academic subjects were found in this cross-sectional analysis. In fact, 2 out of 20 subjects were significantly worse after the change and also Regents exam scores decreased significantly.

**Wahlstrom (2002)^64^** investigated the effect of later SSTs in 7 high schools in Minneapolis, Minnesota, USA, for 3 years before and after the change from a 7:15h to an 8:40h-start. The study analysed objective letters grades and found small improvements that were not statistically significant. However, no actual numbers (or the letter grade scale), nor any statistical test were reported.

#### Cross-sectional studies

**Groen and Pabilonia (2019)^58^** studied N_students_=1200 from a sample of 790 U.S. high schools and reported that a 1h-delay in high school start times was associated with significantly increased reading scores (but not math scores) by 0.16 SD for females, while no significant effect was found for males. The authors used OLS models, including many covariates (individual, family, high school, and community characteristics) that were added sequentially to the models. Data were collected for 2 years, sampled once per year.

**Hinrichs (2011)^54^** found no association between SSTs and ACT scores (N_students_> 196,000) after a delay of 85 minutes from 7:15 to 8:40 AM in 73 schools in Minneapolis, USA, when accounting for various student-level and district level covariates and the length of the school day using OLS regression models (9 years of data). In a similar analysis, the author also found no effect on Kansas assessment scores in reading, maths, science, and social disciplines including all public high schools in Kansas (1,666 schools; up to 5 years of data). In another sample of 75 schools in 19 districts in Virginia, USA, again no association was found between delayed SST and test scores in standardised end-of-course exams (8 years of data).

**Bastian and Fuller (2018)^59^** analysed 4 years of data from N_students_>770,000 in 410 high schools in North Carolina, USA, of which 23 changed their start times (9 schools by ≥30 min). The authors tested both the influence of a linear SST delay per 1h and a categorised school start depending on actual start time i) on overall and 1^st^ period course grades, ii) standardised end-of-course exams, and iii) ACT scores. Linear regression models adjusting for several student and school-level covariates showed that only a start at 8:30h or later was associated with significant improvements of 0.05 SD in 1^st^ period course grades. Importantly, this was one of the only studies that particularly focused on specific subgroups of students: especially low-performers, students with a minority background and with a low SES benefitted from later starts (0.05-0.07 SD in course grades and up to 0.28 SD in ACT composite scores per 1h).

**Dunster *et al*. (2018)^60^** reported results from a cross-sectional pre-post comparison of a one-semester biology course grade from 2 high schools in Seattle, USA, which delayed their starts from 7:50h to 8:45h. Median grade was 77.5% in the year before the delay and 82% in year after the delay (N_students_=178, ~7-month exposure). In logistic regression, where *grade* was used as a predictor variable (not outcome), an increase in grades significantly increased the odds of a student stemming from the after-delay-cohort rather than from the before-delay-cohort (quantitative results not reported; covariates: school, mood, chronotype, sleepiness and sleep offset time on schooldays). Interpretation of the results is limited by the use of grades as independent variable and adjustment for sleepiness and sleep offset in the analysis.

**Milić *et al*. (2014)^53^** analysed in a one-off assessment the final semester grades of 4 Croatian schools (grammar and vocational schools) with morning and afternoon schedules: 2 schools followed early schedules (alternating between 7:00h and 13:00h), while 2 schools had later schedules (8:00h and 14:00h). Based on the sample of N_students_=821 and Mann-Whitney Test (no covariates), it was concluded that students attending the early schedules got significantly better grades (72.0% vs 65.6% in the later-scheduled schools). An extra caveat of the unadjusted analysis is that the sample in the early scheduled schools consisted of three times more boys and grades often associate with gender (see e.g. Fig 3).

**Wolfson *et al*. (2007)^61^** compared the average fall-quarter grade (0-100%) of a total of N_students_=205 attending either an early middle school (starting at 7:15h) or later middle school (starting at 8:37 AM) in New England, USA. MANOVA results with school, grade and gender as covariates indicated that, after half a year of exposure, 8^th^ graders in the later school obtained significantly better objectively reported grades than their early school peers (83.79% vs 76.85%) while no difference was found for 7^th^ graders.

**Lewin *et al*. (2017)^69^** compared 26 middle schools (unknown location) clustered into 3 groups depending on their SSTs (earliest, early, late). The authors obtained self-reported grades (“mainly As”, “mainly Bs”, “mainly Cs”, “mainly Ds/Fs”) and sleep duration from N_students_>32,000 in 3 years. Path-analyses with probit regression with grades as outcome, sleep duration as mediator and inclusion of several covariates showed significantly better grade estimates in the late group compared to the earliest group but not the early group. This effect was in a similar magnitude as that of gender (females better than males) and ethnicity (non-whites worse than whites). Free lunch status as a proxy for SES had the greatest predictive value for grades, while the influence of sleep duration as a mediator was smaller but still significant.

**Wahlstrom *et al*. (1997)^62^** compared middle and high schools in three districts in Minnesota, USA, of which District A delayed its start time to 8:30 and Districts B and C stayed with their earlier starts of 7:25h and 7:15h, respectively. The study reports that mean self-reported grades in high schools in district A were highest compared to the other 2 districts, however the scale, statistical analyses and the use of covariates were not reported. Results for middle schools (7-8^th^ graders) were comparable but again no statistics were given and absolute differences were marginal.

**Kelley et al. (2017)^68^** followed an English high school over 4 years across two changes in SST (A-B-A design): from the standard school start at 8:50h in year 0, to a delayed start at 10:00h for two years, and for another year back to the original start time. For each year, national examination results (GCSE exams, achievement over the past 2 years plus final examination) of a different cohort of students was assessed (N_students_>2,000). As the only study in our selection, the study analysed the achievement of the cohorts not only in comparison to each other but also in comparison to a national benchmark and to an indicator of predicted progress (value-added prediction) based on student cohorts’ past achievements. Based on t-tests (no covariates), the study found that delayed SST for two years were significantly associated with an increased percentage of students making good academic progress (*i.e*., achieving ≥5 GCSE grades of ≥C in English, math and ≥3 other subjects) (from 34% in year 1 to 52% in year 3) and with a 12-percentage point increase in the value-added number of students. Both improvements were partly reversed after return to the earlier start time.

**Wahlstrom *et al*. (2014)^63^** analysed self-reported grades, objectively reported GPAs, and standardised test scores (state-wide achievement tests or PLAN) from 9-12^th^ graders after a high school start delay from 7:35h-7:50h to 8:00h-8:55h in Minnesota, Colorado, and Wyoming, USA. The study yielded mixed and mostly not-significant effects using t-tests and correlations without considering covariates.

### Risk of bias assessment

To judge the evidence quality of the included studies, we performed a risk of bias assessment (Tab 1). Overall, since none of the studies were RCTs, selection bias was high by definition for all studies. Furthermore, in many studies, basic reporting standards were only partially met (reporting bias), blinding was a high concern in over half of the studies (performance bias), and appropriate statistical models that control for confounders were not used in 7 out of 21 studies. This meant that over half of the studies stayed below 75% of the good-evidence-score within their respective study design category. Therefore, the quality of the evidence can be deemed only moderate.

On the positive side, especially the longitudinal studies with a control group showed a high evidence quality with 2^56,65^ out of 6 studies reaching at least a 75%-score and 3 more studies^48,55,67^ >50%. Two studies^48,67^ could have improved their score to 75% simply by ensuring sufficient reporting of outcomes and statistical analyses. Furthermore, all included studies had appropriately large sample sizes (and/or high resolution) and were therefore very likely suited to detect a true effect (sufficient statistical power).

### Magnitude of effects

Given that a meta-analysis was not indicated because of the great variation in outcome measures, study designs and analyses, we identified a subgroup of studies that provided standardised beta coefficients to compare the magnitude of grade or score changes across studies and with those of covariates. The statistically significant beta coefficients of the identified 8 studies are plotted in Figure 3a. Overall, the magnitude of the influence of school start times is smaller than SES or ethnical/racial background. In line with this, studies from Edwards^55^ and Bastian and Fuller^59^ demonstrated that it was the disadvantaged and minority students that particularly benefitted from later starts. Importantly, Figure 3a purports a biased picture towards positive results of school start times on achievement given the selection for significant standardised coefficients. Figure 3b puts findings from 3a into perspective with all included studies and paints a very different picture.

## Discussion

Chronic sleep restriction in teenagers has become a serious health concern worldwide^e.g. 8,70^. The widespread sleep restriction is largely a result of the conflict between the late sleep times typical of adolescence and the early SSTs imposed by society^e.g. 3,71,72^. Delaying school start times has the great potential of improving cognitive functioning, physical health and well-being of students mediated by improving sleep (as reviewed elsewhere^16,25,31^) with possibly relatively little costs^73,74^. But does a delay in SSTs also translate into improved academic achievement in middle and high school students? Our systematic literature search identified 21 studies that investigated whether SSTs have any systematic effect on course grades or (standardised test) scores. The analyses revealed that about half of the studies did not find any effect (neither positive nor negative) of delaying school times on academic performance, while the other half found either mixed or positive results. Just one study found better grades associated with earlier SSTs^53^. Given the high risk of biases observed in most of the studies and the great heterogeneity in school settings, there is a need for more high-quality evidence to draw sound conclusions (see Fig. 4 for suggested improvements).

**Fig. 4.**
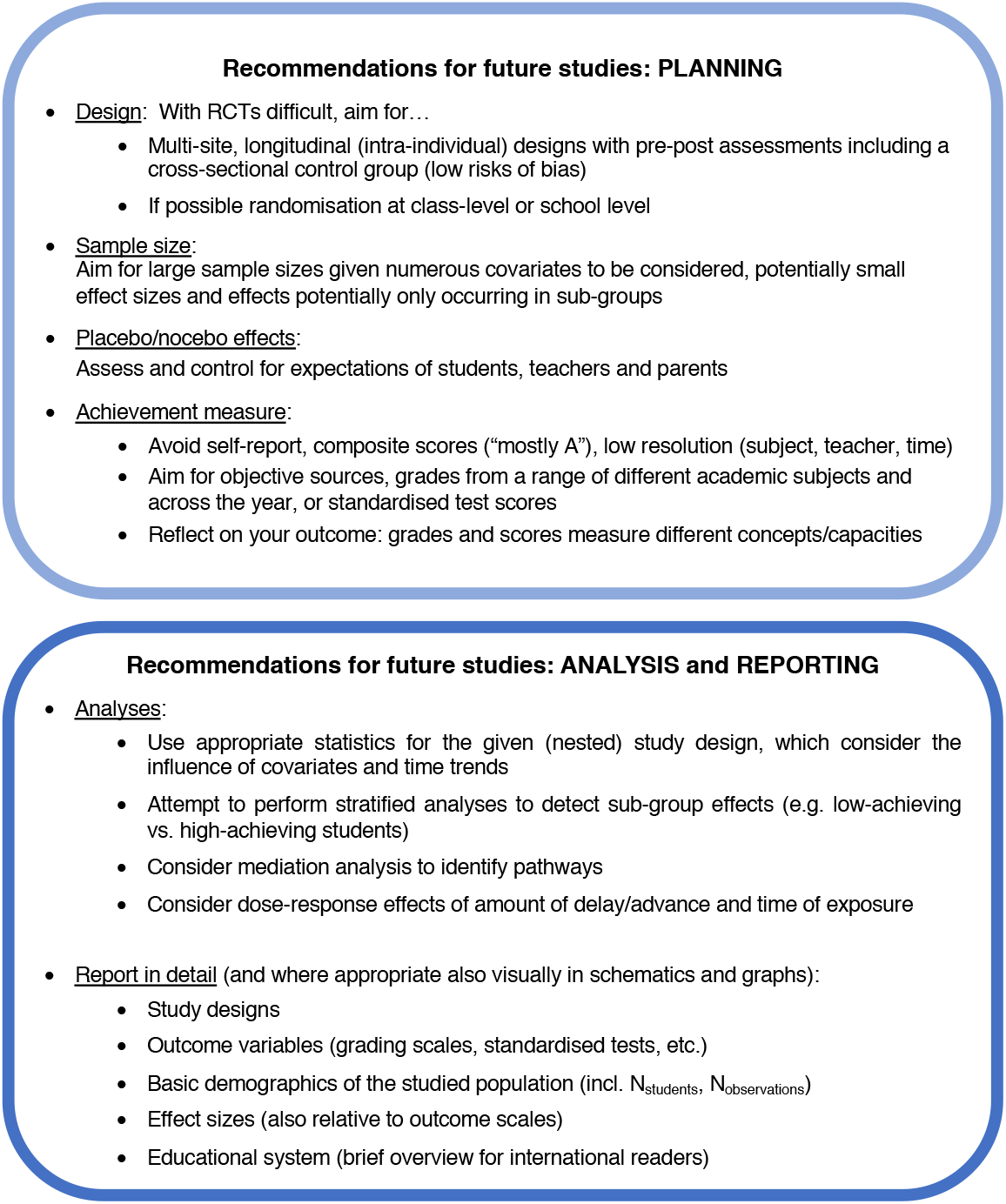
Suggestions for future studies.

### Methodological considerations

Our systematic risk of bias assessment showed that the evidence level was mostly moderate (only 5 out of 21 studies achieved a score of ≥75% within their category). Specifically, we did not identify any randomised controlled trials, which is not surprising considering the circumstances of educational research and the hesitation of many schools to participate in such complex and time-consuming study designs^75^. In many studies, basic reporting standards were only partially met, blinding was a high concern in over half of the studies (i.e. high performance bias), and appropriate statistical models which control for confounders were not used in 7 out of 21 studies.

The study design we identified as one of the most commonly used was longitudinal studies with a pre-post design that followed a specific cohort of students over time i) including a control group that did not change start times, and ii) without a control group. A second common design was cross-sectional studies that compared different, independent groups of students (either at one specific time point or over several years) with varying start times. Studies that performed best in the risk of bias assessment were mostly longitudinal studies with a control group, a large sample size and with appropriate and advanced statistical analyses that controlled for possible confounders.

### Well-designed studies also reveal no clear picture

What do studies with low risk (*i.e*., a ≥75% good-evidence-score) conclude about the influence of SSTs on academic achievement? Lenard *et al*. (2020)^56^ found that advancing SSTs by 40 minutes did not affect ACT scores, while Jung (2018)^65^ showed that delaying start times by 40-60 min also did not affect grades when personal covariates were controlled for. If studies with a good-evidence-score of 50% are also considered, the picture is more complex: two studies report small gains in math and reading^48,55^, and one reports small effects on math but not on Korean nor English^67^. Three cross-sectional studies also achieved a good evidence score of over 75%^54,58,59^. The associations found between SSTs and academic achievement again did not point in one direction: Groen and Pabilonia (2019) considered a range of different start times and reported small increases on the Woodcock-Test but only for females and reading^58^, while Hinrichs did not find any positive association of a delay of 85 min on either ACT scores, Kansas assessment scores, or end of course exams^54^. Bastian and Fuller (2018) reported that a 8:30h or later start was necessary for positive associations with 1^st^ period grades^59^. Furthermore, the authors showed that especially lower achievers, minority students and students with a low SES benefit from later starts. In summary, good evidence studies report either no, relatively small, or not generalisable effects of changing SSTs.

### Do results for course grades and standardised test scores differ?

Since course grades and standardised scores possibly measure different underlying skills and knowledge, they might also differ in their sensitivity to SST changes. For instance, standardised test scores seem to be sensitive enough to reflect effects of other school policies, *e.g*. reducing classroom size^76^ or racial segregation^77^. However, general test scores might be less sensitive to acute changes in SSTs because they measure the accumulated knowledge over several schooling years^56^ and are taken predominantly by high-achieving students, who are prone to ceiling effects. Moreover, they are often scheduled in the morning^54^ and therefore confounded by time-of-day effects on attention and fluid intelligence (*e.g*. logic, reasoning, problem solving)^56,78–80^. Moreover, in the case of ACT or PLAN scores, tests are usually only taken by high-achieving students applying for admission to college – a specific student population which is prone to ceiling effects, making these students less likely to benefit from later SSTs compared to lower-achieving students as two other studies also confirmed^55,59^.

Course grades, on the contrary, derive from exams taken by all students. If collected with high temporal resolution (*i.e*. more than once per year), they are potentially more sensitive to acute SSTs changes and less influenced by time-of-day effects if distributed evenly across the day. However, grades might be more influenced by certain student characteristics, such as conscientiousness or perseverance^81^. A “teacher bias” could particularly influence the results of interventional studies if not controlled for. Altogether, both standardised test scores and course grades have their pros and cons, which might be the reason why no clear answer emerges even when results are grouped by outcomes: there was no tendency or differential effect on either objective test scores (2 positive^55,68^, 3 null findings^54,56,57^, and 4 mixed results^58,59,63,67^) or objective grades (2 positive^48,60^, 4 null findings^49,57,64,65^ and 2 mixed results^59,61^) or self-reported grades (2 positive^62,69^, 3 null findings^51,52,66^, and 1 each for mixed^63^ and negative^53^).

### Considerations of power and dose

An alternative explanation for these mixed results could be a lack of statistical power. However, almost all studies had very large sample sizes or number of observations and were able to detect other influences such as gender differences and achievement gaps between whites and non-whites. The effect sizes of these factors tended to be of larger magnitude than effect sizes for changes in school starts (Fig. 3a).

Another interesting consideration is that effects of changed SSTs on achievement might not be linear. When exactly should schools start? How much should schools delay their start times? How long do students need to be exposed to later starts until effects become visible? These are important practical questions that are, however, difficult to answer. Intuitively, one would expect that small delays are not enough to produce robust effects. However, it is not clear whether further delays would be beneficial or even harmful. Hinrichs^54^ tried to model this hypothesis using spline regressions but found no clear answer. Furthermore, the latest start time in the studies reviewed here was 10:00h and the largest delay was 135 min (Fig. 2e). Despite a great variation in delays and SSTs, we were not able to detect any clear dose response curve, *i.e*. positive effects only appearing with the largest delay. Further studies should clarify this question. Nevertheless, the American Association of Pediatrics recommends to start schools not earlier than 8:30h^82^, which is supported by Bastian and Fuller^59^ who found that only when school started at 8:30h or later, significant positive effects were detected on 1^st^ period grades, although overall grades were unaffected.

A second consideration about dose is how long the school has already operated in a delayed system – the longer the delay has been in place, the longer students were exposed. Several studies analysed time trends for several years before and after a change but no unifying results emerge from these studies.

### Factors influencing academic achievement

A very likely reason for inconclusive results derives from the many variables affecting course grades and test scores. Whether these variables are assessed, considered, and controlled for can drastically change the conclusions of a study. These influences range from student-level factors (*e.g*. chronotype^83^, ethnic or racial background^59^, conscientiousness^81^ or prior knowledge^84^) to family-level factors (*e.g*. parental involvement^85^, parental education^65^, or SES^86^), and to classroom- and school-level factors (*e.g*. classroom size^76^ and atmosphere^84^, teacher quality^87^). Indeed, we also observed here that SES and race/ethnicity influence achievement (Fig. 3a). Moreover, there are sleep variables, such as sleep duration and daytime sleepiness that play an important role for health, cognition and learning and are often connected to demographic variables, such that students with difficult social backgrounds are also prone to reduced and poorer sleep than their more advantaged peers^88,89^. It is therefore likely that SST delays potentially only translate into meaningful grade or test score benefits in a specific subset of students. Stratified analyses could answer this question but have rarely been done (for notable exceptions see^55,59^ which confirm such tendencies). In general, reflecting on confounders, their influence on academic achievement and on how they might also be affected by changes in SSTs is important for designing future studies.

### Limitations of the review

Although an extensive search across different databases was carried out, an incomplete retrieval of all published articles on the topic cannot be excluded. A total of 21 studies were included, which is far more than in previous reviews (2-12 included studies). We also chose to report grey literature to reduce a possible publication bias in favour of positive results. Previous reviews^16^ decided otherwise to ensure a good quality of the findings reported. However, the included risk of bias assessment allowed for critical reporting of both peer and non-peer-reviewed articles. Since the studied population was restricted to middle and high school students, several studies which used valuable randomisation at the class-level had to be excluded because they included college students (for a review see^24^). However, life-style and sleep characteristics widely differ between high school and college students, which is why we focused only on adolescents. We included middle schools, since sleep changes tend to start with the onset of puberty ^90,91^.

### Final Conclusions

Our systematic research and analysis of the literature shows that the current evidence does not allow to draw sound conclusions as to whether delaying SSTs improves or is associated with increased achievement at the grade and test score level across all students. This is mostly due to the heterogeneity in school settings and the vast differences between studies with regards to study design, quality and chosen outcome measure and consequently a lack of generalisability of individual study results that also prevented conducting a meta-analysis (see Fig. 4 for suggested improvements). Importantly, as much as course grades and test scores do not *systematically* or greatly improve across the majority of studies, all included studies (except for one) showed no worsening after a SST delay. This means that SSTs could be delayed, while academic achievement is maintained at the same level (or improved in sub-groups or individuals) and possibly achieved with less cognitive effort or time spent on studying and homework (students are likely better rested and therefore cognitively more capable and efficient). In combination with other reported positive outcomes on sleep, daytime sleepiness, mood and motivation, computer gaming, attendance rates, or tardies and suspensions^e.g.14,15,24,25,33^, this remains a valid argument in favour of delaying SSTs.

## Data Availability

NA since this was a systematic review. Authors can provide more details regarding literature search upon request.

## Acknowledgements

We thank all contacted authors who replied and helped us to report their findings as accurately as possible. AMB thanks the Graduate School for Systemic Neurosciences, Munich, for financial support.

## Author contributions (CRedIT Taxonomy)

Conceptualisation: AMB, GZ, ECW

Methodology: AMB, GZ, ECW, KM

Investigation: AMB, GZ

Data curation: AMB, GZ

Formal analysis: AMB, GZ

Validation: ECW, KM

Supervision: ECW

Visualization: AMB, GZ

Writing – original draft: AMB, GZ

Writing – review and editing: AMB, GZ, ECW, KM

## Supplementary information

**Tab S1.**
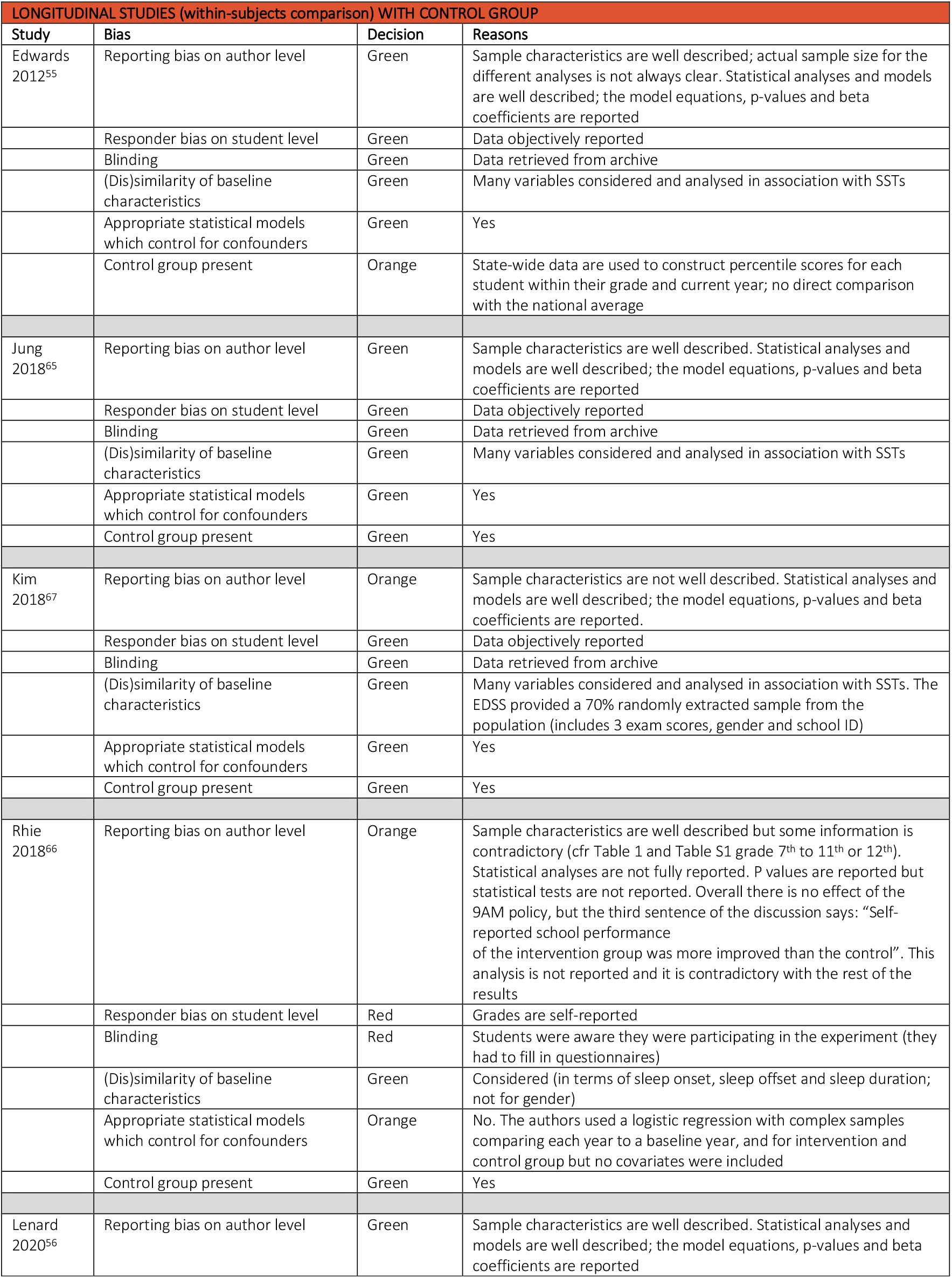
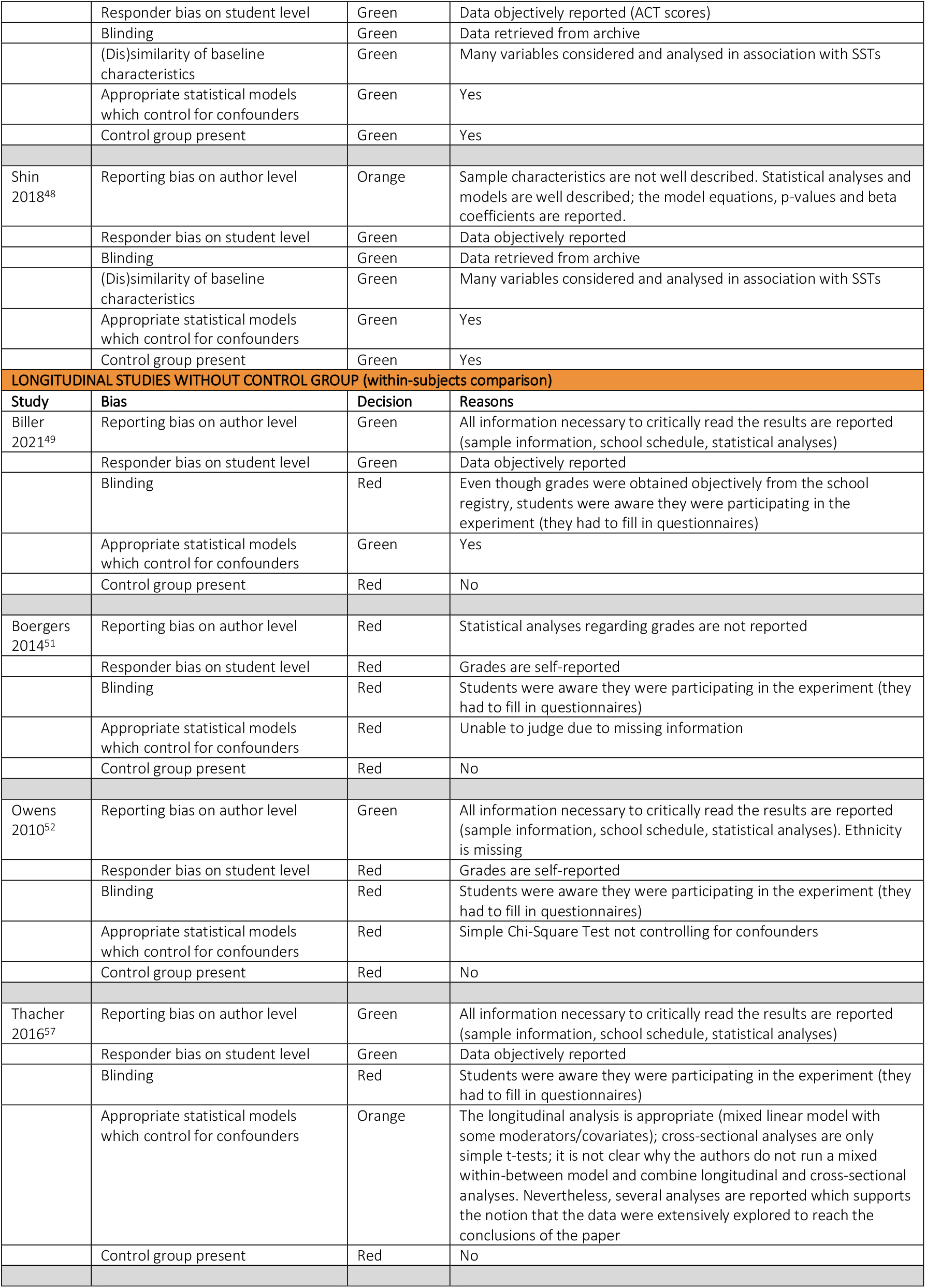
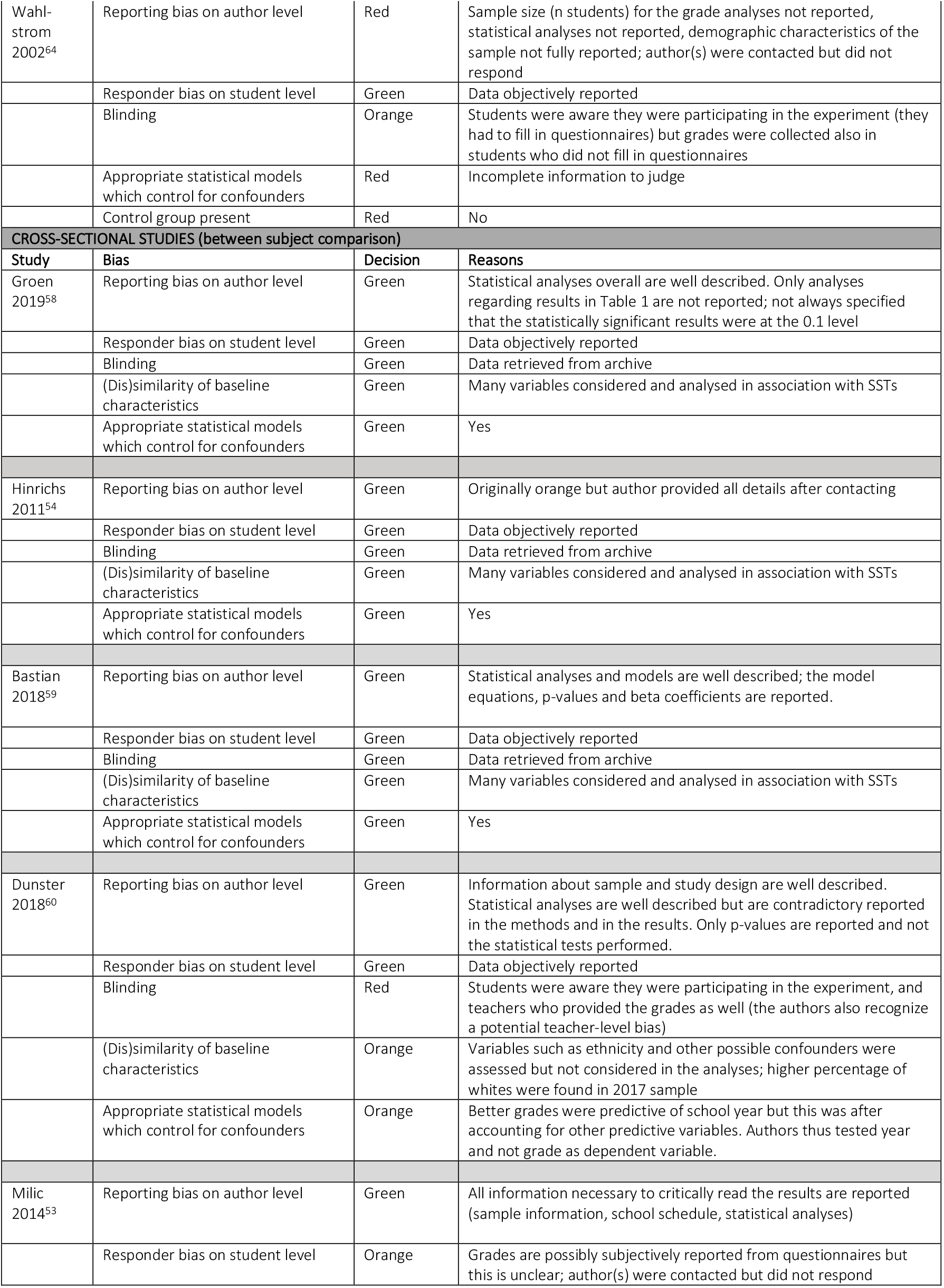
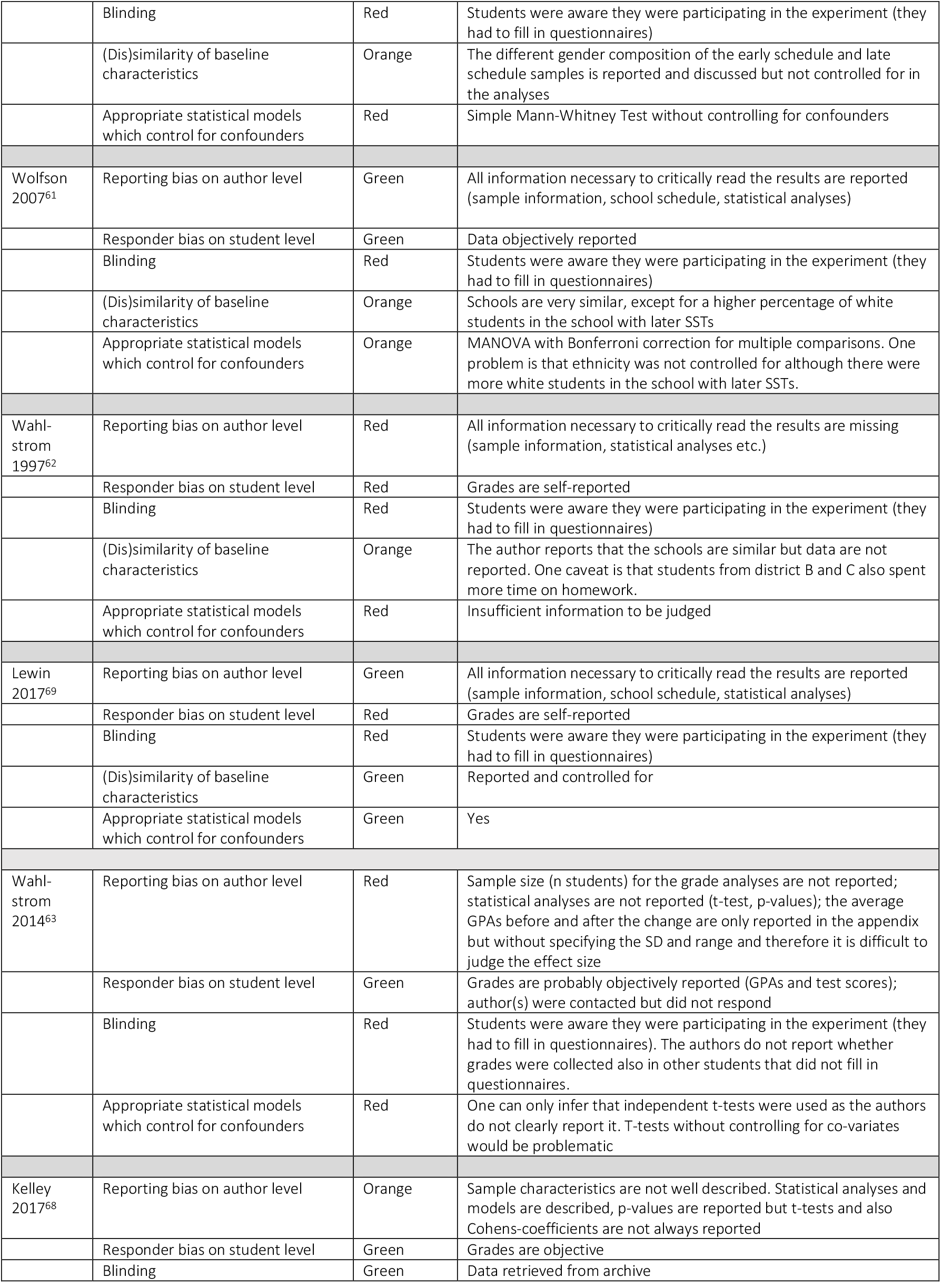
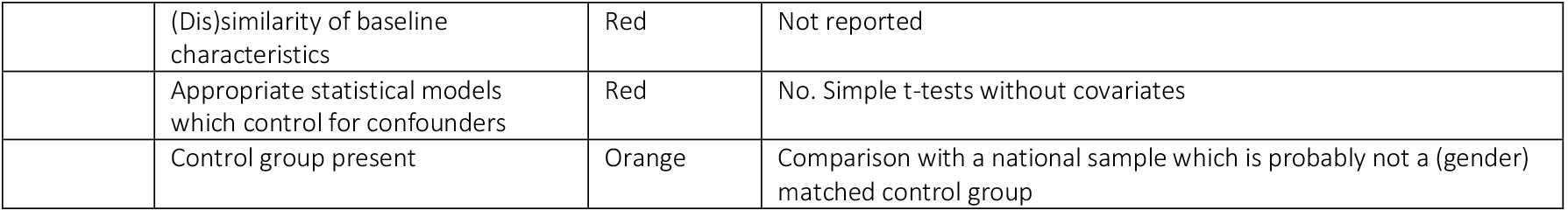
Protocol detailing reasons for risk of bias assessment decisions of Tab. 1.

